# Mitigating Automation Bias in Physician-LLM Diagnostic Reasoning Using Behavioral Nudges: A Randomized Controlled Trial

**DOI:** 10.64898/2026.06.01.26354596

**Authors:** Ihsan Ayyub Qazi, Ayesha Ali, Asad Ullah Khawaja, Muhammad Junaid Akhtar, Ali Zafar Sheikh, Muhammad Hamad Alizai

**Affiliations:** Department of Computer Science, Lahore University of Management Sciences, Lahore, Pakistan; Department of Economics, Lahore University of Management Sciences, Lahore, Pakistan; King Edward Medical University, Lahore, Pakistan; Lahore General Hospital, Lahore, Pakistan; Children’s Hospital, Lahore, Pakistan

## Abstract

As large language models (LLMs) enter clinical workflows, automation bias, the uncritical acceptance of automated output, poses a patient-safety risk. Optimal physician-AI collaboration requires trust calibration, matching scrutiny to LLM recommendation accuracy. We report a randomized trial evaluating a behavioral nudge to mitigate automation bias. Seventy-two AI-trained physicians were randomized to evaluate six vignettes alongside ChatGPT-5.1 recommendations, consulted at each physician’s discretion; three contained deliberate, clinically significant errors. The treatment arm received a dual-component nudge: an anchoring cue reporting ChatGPT’s benchmark accuracy to calibrate expectations, and a case-specific, selective-attention cue; a numeric accuracy rating and color-coded traffic light, derived from the mean of three distinct-family LLMs. The control group saw recommendations alone; blinded reviewers scored diagnostic reasoning against an expert rubric. The treatment group scored significantly higher (mean difference, 7.6 percentage-points; 95% CI, 1.4-13.9; *P*=0.016) than the control, suggesting a scalable strategy to preserve clinical judgment in LLM-assisted care. ClinicalTrials.gov registration: NCT07328815.

---

Diagnostic errors remain a major source of preventable harm, contributing to an estimated 795,000 deaths or permanent disabilities each year in the United States and between 5.7-8.4 million excess deaths annually in low- and middle-income countries.^1-5^ Most of these errors arise from breakdowns in clinical judgment; premature anchoring on a narrow differential, misinterpreted test results, delayed specialist referral,^6-8^ and precisely the reasoning that large language models (LLMs) are now being adopted to augment.^9-13^ But these tools carry a new hazard: LLMs “hallucinate,” producing fluent, coherent, yet factually incorrect output,^14-16^ and when physicians consult them, hallucination collides with automation bias; clinician’s well-documented tendency to favor an automated system’s recommendation even when it is wrong.^17-21^ This susceptibility is not a marginal lapse but a structural feature of clinical work: time pressure in high-volume settings, incentives that reward throughput over deliberation, and cognitive fatigue across long shifts erode the capacity for sustained scrutiny, while diffusion of responsibility, overconfidence in technology, and cognitive offloading further predispose clinicians to accept AI-generated output uncritically.

The risks are not hypothetical: even flagship models still fabricate clinically significant content and can be manipulated into issuing dangerous recommendations,^22,23^ physicians given inaccurate decision support make significantly worse diagnoses,^24^ and LLM tools themselves shift toward less appropriate care under anchoring cues.^25^ In a recent randomized trial, physicians who had completed a 20-hour AI-literacy curriculum and retained full discretion over LLM use nonetheless showed significantly degraded diagnostic reasoning when ChatGPT-4o recommendations contained errors.^26^ Critically, AI-literacy training alone, currently the dominant mitigation strategy endorsed by health-system leadership,^27-30^ was insufficient to protect physicians from plausible-sounding but incorrect recommendations. This establishes automation bias as a robust phenomenon in physician-LLM collaboration and raises an urgent question: beyond pre-encounter education, what point-of-care interventions can recalibrate physician reliance, curbing deference to incorrect recommendations while preserving the value of correct ones?

A reflexive response to the documented risks of LLM hallucinations is to instruct physicians to distrust AI output by default.^31^ This approach is both impractical and counterproductive. Studies show that LLM consultation, when recommendations are correct, improves diagnostic accuracy by margins on the order of 18 percentage points,^32^ and on-demand consultation models that preserve clinician autonomy are being rapidly adopted across emergency medicine, primary care, and radiology workflows.^10-12,33-35^ The goal, therefore, is not uniform skepticism but trust calibration: matching the depth of physician scrutiny to the likelihood that a given LLM recommendation is correct. Calibration at the point of care requires solving two interlinked problems. First, physicians need a stable prior, an evidence-based expectation of how often the LLM is right on tasks of this kind, to anchor their evaluation, since clinicians’ subjective estimates of model accuracy are poorly calibrated.^17-21^ Model-generated confidence signals are themselves unreliable, with documented overconfidence on incorrect outputs.^36^ Second, they need a case-specific signal flagging which recommendations warrant deeper analytical engagement, since the narrative sophistication of LLM output can otherwise suppress the kind of effortful System 2 reasoning needed to catch subtle errors.^37^ Both signals must be lightweight enough to integrate into existing clinical workflows without imposing the cognitive cost that would defeat the purpose of consulting AI. Prior human-computer interaction research has tested two main interventions for AI overreliance, model self-explanations and workflow-disrupting cognitive forcing functions, but each falls short: explanations are typically processed heuristically rather than substantively, while forcing functions reduce overreliance only at the cost of uniform friction that users resist.^37^ To our knowledge, no prospective trial has tested whether a behavioral intervention designed around these two principles, anchoring physicians to an evidence-based prior on LLM accuracy and selectively cueing analytical scrutiny on high-uncertainty recommendations, can mitigate automation bias in physician-LLM diagnostic collaboration.

Here we report a randomized controlled trial evaluating a dual-component behavioral nudge intended to support trust calibration during voluntary LLM consultation. Seventy-two physicians, all of whom completed standardized AI-literacy training, were randomized to treatment or control arms and evaluated six clinical vignettes accompanied by ChatGPT-5.1 diagnostic recommendations, three of which contained deliberately introduced, subtle, and clinically significant errors (432 cases total). Vignette sequencing and error placement were randomized to control for order effects and prevent pattern recognition, and LLM consultation remained at physicians’ discretion. The treatment arm received a nudge comprising two transparency-based cues alongside each recommendation: an anchoring cue reporting ChatGPT’s diagnostic accuracy on standard benchmarks, intended to calibrate baseline expectations, and a selective-attention cue in the form of a case-specific signal that displays the mean of three confidence ratings independently assigned by three state-of-the-art LLMs from distinct model families (Claude Sonnet 4.5, Gemini 2.5 Pro Thinking, and GPT-4o), together with a color-coded traffic-light signal derived from that mean score (red below baseline, orange above baseline but below 100%, green at 100%), intended to prompt System 2 engagement on recommendations with high uncertainty. The control arm received the same recommendations without the nudge. Diagnostic reasoning was scored by blinded reviewers against an expert-developed rubric. We hypothesized that the nudge would attenuate automation bias without erasing the benefits of LLM consultation on error-free cases, providing a scalable, workflow-compatible alternative to education-only mitigation. Our findings demonstrate that a lightweight, transparency-based intervention combining calibrated performance anchoring with ensemble-derived confidence signaling can meaningfully preserve clinical judgment in physician-AI collaboration, a result with direct implications for the safe deployment of LLM-assisted decision support across high-stakes clinical settings.

## Results

Seventy-two licensed physicians were recruited from January 1 to May 22, 2026. 36 were assigned to the intervention group and 36 to the control group (Fig. 1). Of these, 37 participants (51%) completed virtual study sessions and 35 (49%) completed in-person sessions. Participants represented 47 medical institutions across Pakistan. The LLM recommendation panel was opened by participants in 91.7% of intervention cases and 93.5% of control cases, a difference that was not statistically significant (*P*=0.46). The median years in practice was 8.0 years (interquartile range (IQR) = 3.0 to 14.0 years). Baseline participant characteristics are summarized in Table 1.

**Table 1.** Baseline Participant Characteristics.

| Table 1. Baseline Participant Characteristics |  |  |  |  |
| --- | --- | --- | --- | --- |
|  | Participants, No. (%) |  |  |  |
| Participant Characteristic | Overall (n=72) | Physicians plus conventional resources plus nudge (n=36) | Physicians plus conventional resources only (n=36) | P value |
| Gender |  |  |  |  |
| Male | 25 (35) | 11 (31) | 14 (39) | 0.62 |
| Female | 47 (65) | 25 (69) | 22 (61) |  |
| Specialty |  |  |  |  |
| Physician (primary care & medical specialties)<sup>a</sup> | 36 (50) | 18 (50) | 18 (50) | 0.59 |
| Surgeon<sup>b</sup> | 22 (31) | 13 (36) | 9 (25) |  |
| Diagnostic/Support Specialties<sup>c</sup> | 3 (4) | 1 (3) | 2 (6) |  |
| Other/Non-Clinical<sup>d</sup> | 11 (15) | 4 (11) | 7 (19) |  |
| Years in practice |  |  |  |  |
| Mean (SD) | 9.1 (7.9) | 9.8 (7.9) | 8.3 (7.8) | 0.42 |
| Median (IQR) | 8.0 (3.0-13.0) | 8.0 (3.0-14.0) | 6.0 (2.8-12.0) | 0.34 |
| LLM experience |  |  |  |  |
| Infrequent Use (less than weekly) | 21 (29) | 13 (36) | 8 (22) | 0.30 |
| Frequent Use (weekly or more) | 51 (71) | 23 (64) | 28 (78) |  |
| Notes: |  |  |  |  |
| Percentages in parentheses may not sum to 100% on account of rounding.<br>Participant characteristics (gender, specialty, LLM experience) were compared between groups using the chi-square test ($\chi^2$) test, or Fisher’s exact test when any expected cell count was less than 5. Continuous variables (years in practice) were compared using two-sided Welch’s t-test (unequal variances) for the mean and the Mann-Whitney U-test for the median. Reported P values are two-sided; no adjustments were made for multiple comparisons; LLM, large language model; and SD, standard deviation.<br>Self-reported LLM experience was dichotomized for analysis. Participants were grouped into ‘Frequent Use’ (less than once per week use) and ‘Infrequent Use’ (at least once per week use).<br>Specialty details.<br><sup>a</sup> Includes primary care fields (family medicine/general practice, internal medicine, pediatrics) as well as medical specialties (cardiology, neurology, gastroenterology, emergency medicine, psychiatry).<br><sup>b</sup> Includes general surgery, obstetrics/gynecology, urology; ophthalmology; otolaryngology (ENT); breast surgery; hepato-pancreato-biliary and liver surgery; pediatric neurosurgery; and oral and maxillofacial surgery.<br><sup>c</sup> Include radiology, pathology, and anesthesiology.<br><sup>d</sup> Includes community medicine/public health, research positions, and house officer |  |  |  |  |

**Figure 1:**
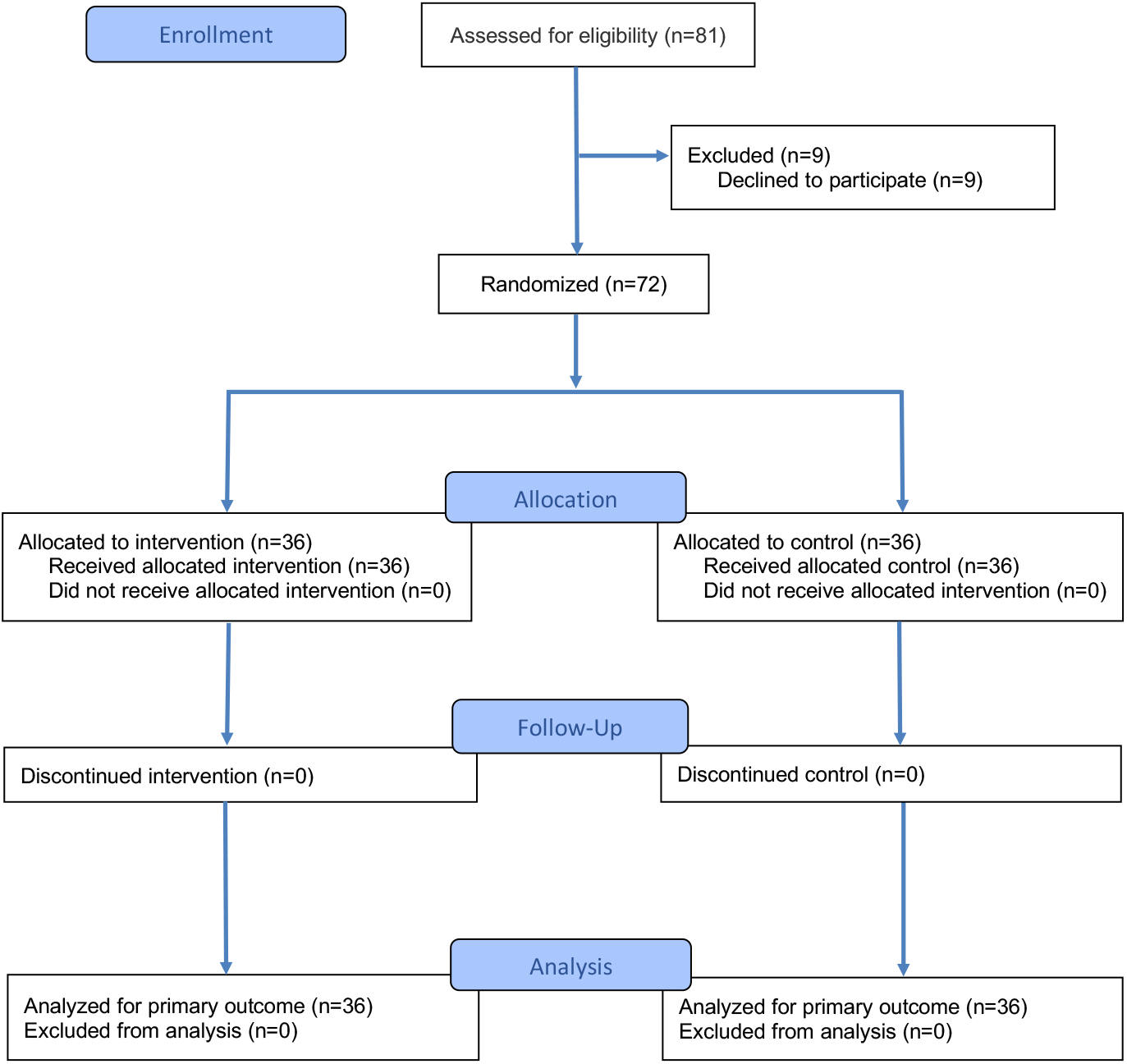
Participant flow. The study included 72 physicians, who were randomized 1:1 to the intervention or control arm (n=36 each), to test whether a behavioral nudge reduces automation bias, the uncritical acceptance of incorrect LLM recommendations, in clinical decision-making. Each participant evaluated six clinical vignettes (432 evaluations total), presented in randomized order, spanning a range of diagnostic difficulty and specialties. Three vignettes contained deliberately introduced reasoning errors in the accompanying LLM recommendations and three contained correct recommendations; vignette content was identical across arms. For every vignette, a fixed, pre-generated diagnostic recommendation from ChatGPT-5.1 was available on a custom web platform but displayed only when the physician chose to view it. Three of the six recommendations contained deliberately introduced reasoning errors and three were correct, with vignette content identical across arms. Control-arm physicians who opted to view the recommendation saw it without additional cues. Intervention-arm physicians who opted to view the recommendation received a dual-mechanism nudge: a baseline-accuracy anchoring cue and case-specific signal that displays the mean of three confidence ratings independently assigned by three state-of-the-art LLMs from distinct model families (Claude Sonnet 4.5, Gemini 2.5 Pro Thinking, and GPT-4o), together with a color-coded traffic-light signal derived from that mean score (red below ChatGPT-5.1’s baseline accuracy; orange, above baseline and below 100%; green, 100% consensus). Physicians in both groups could voluntarily consult the LLM consultation alongside conventional diagnostic resources (e.g., PubMed, Google Search without AI features). The primary outcome was the diagnostic reasoning accuracy score; the secondary outcome was the top-choice diagnosis accuracy score. Responses were scored by blinded reviewers against rubrics developed by three licensed physicians.

### Primary Outcome

A total of 432 cases were completed by the participants (216 in the treatment group, and 216 in the control group). All participants completed each of the six vignettes. The mean (SD) score per case was 82.8% (24.7%) for the treatment group and 75.6% (28.7%) for the control group. The linear mixed effects model showed a difference of 7.6 percentage points (95% CI: 1.4 to 13.9 pp; *P*=0.016) between the treatment and control groups (Table 2 and Fig. 2).

**Table 2.**
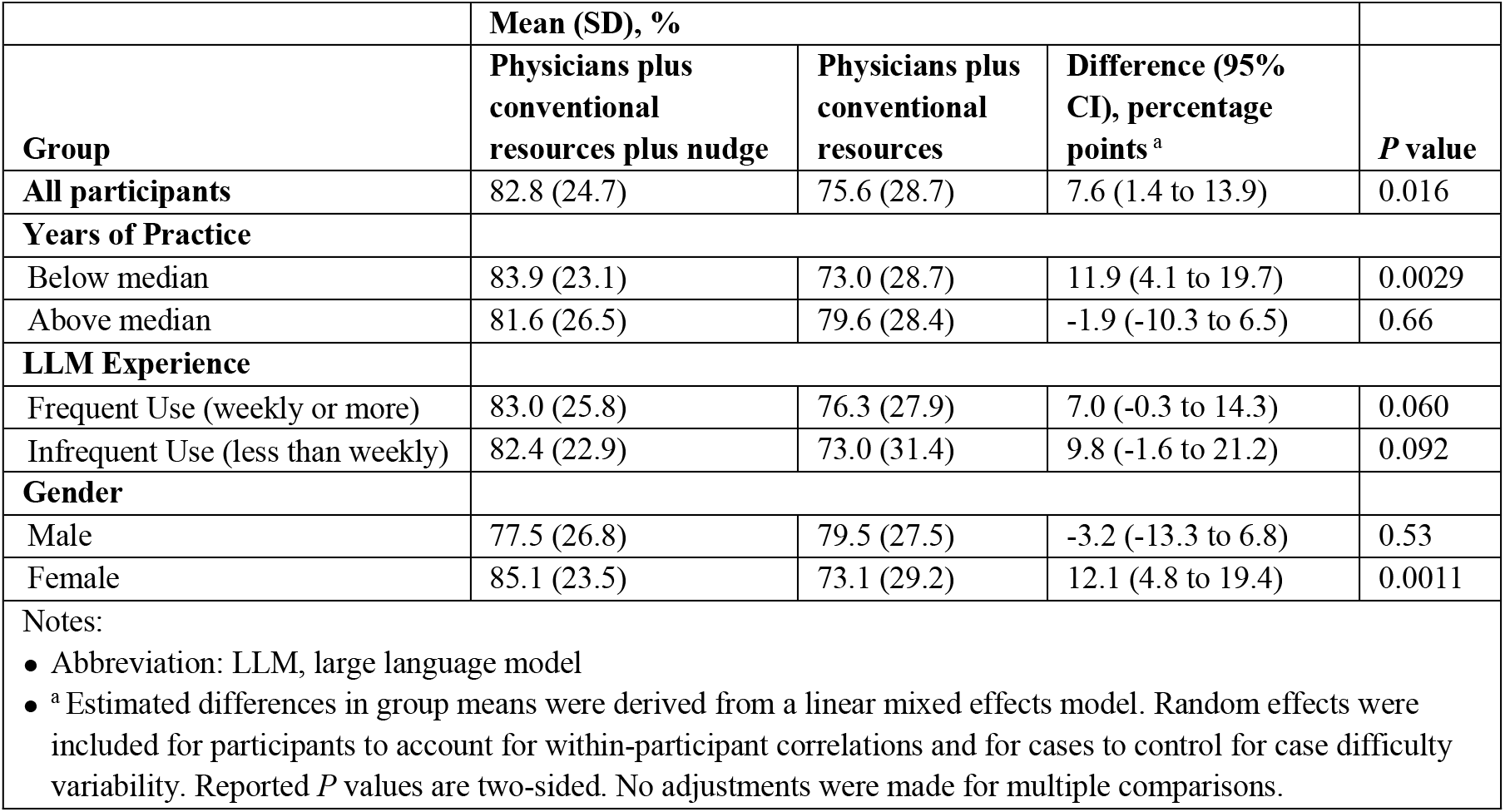
Diagnostic Reasoning Accuracy Outcome.

| **Table 2. Diagnostic Reasoning Accuracy Outcome** |  |  |  |  |
| --- | --- | --- | --- | --- |
|  | **Mean (SD), %** |  |  |  |
| **Group** | **Physicians plus conventional resources plus nudge** | **Physicians plus conventional resources** | **Difference (95% CI), percentage points <sup>a</sup>** | **P value** |
| **All participants** | 82.8 (24.7) | 75.6 (28.7) | 7.6 (1.4 to 13.9) | 0.016 |
| **Years of Practice** |  |  |  |  |
| Below median | 83.9 (23.1) | 73.0 (28.7) | 11.9 (4.1 to 19.7) | 0.0029 |
| Above median | 81.6 (26.5) | 79.6 (28.4) | -1.9 (-10.3 to 6.5) | 0.66 |
| **LLM Experience** |  |  |  |  |
| Frequent Use (weekly or more) | 83.0 (25.8) | 76.3 (27.9) | 7.0 (-0.3 to 14.3) | 0.060 |
| Infrequent Use (less than weekly) | 82.4 (22.9) | 73.0 (31.4) | 9.8 (-1.6 to 21.2) | 0.092 |
| **Gender** |  |  |  |  |
| Male | 77.5 (26.8) | 79.5 (27.5) | -3.2 (-13.3 to 6.8) | 0.53 |
| Female | 85.1 (23.5) | 73.1 (29.2) | 12.1 (4.8 to 19.4) | 0.0011 |
| Notes:<br>• Abbreviation: LLM, large language model<br>• <sup>a</sup> Estimated differences in group means were derived from a linear mixed effects model. Random effects were included for participants to account for within-participant correlations and for cases to control for case difficulty variability. Reported *P* values are two-sided. No adjustments were made for multiple comparisons. |  |  |  |  |

**Figure 2:**
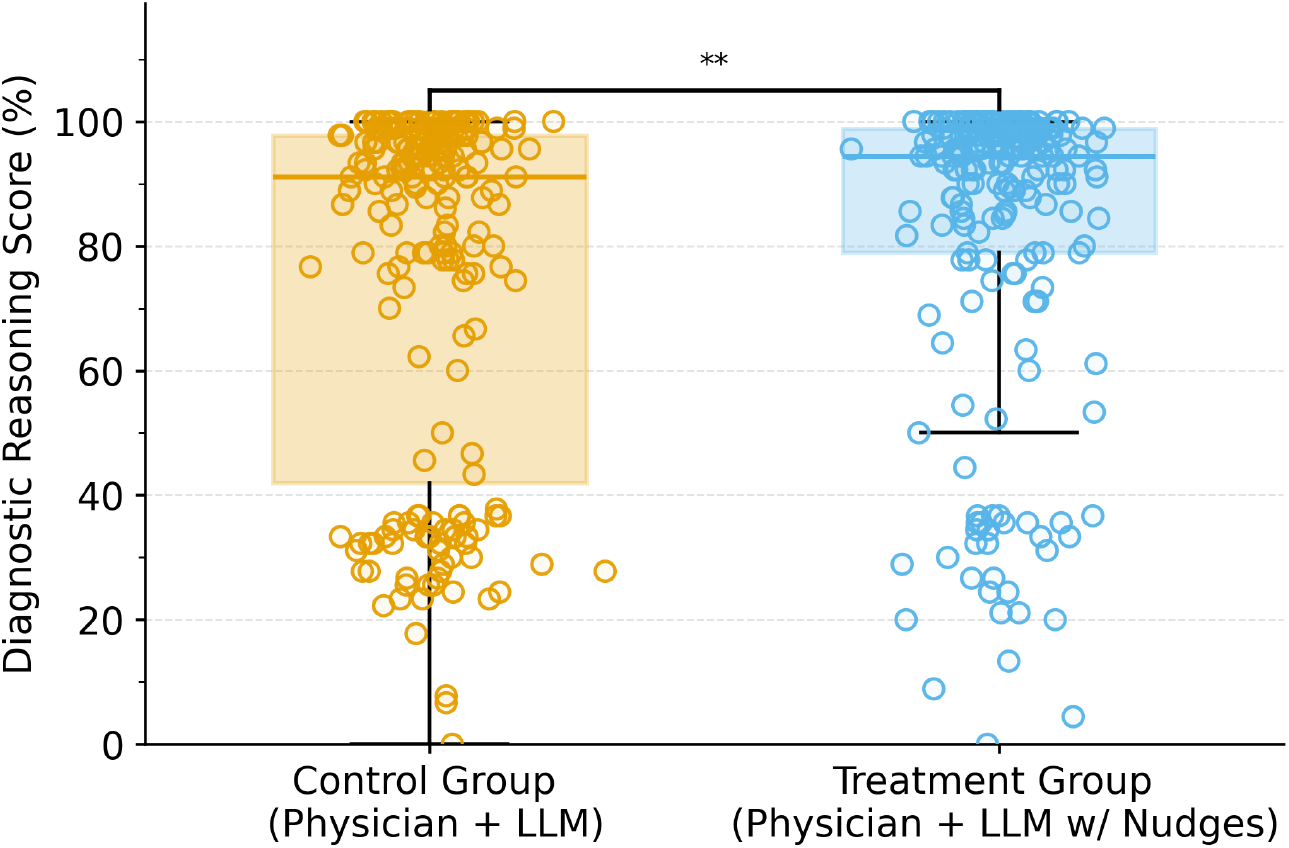
Comparison of the primary outcome for physicians in the treatment group with physicians in the control group (diagnostic reasoning score standardized to 0-100). Seventy-two physicians were randomized 1:1 and completed 432 cases (216 in the treatment group, 216 in the control group). The center of the box plot represents the median, with the boundaries representing the first and third quartiles. The whiskers represent the furthest data points from the center within 1.5 times the IQR. Individual data points show case-level scores. The treatment group demonstrated significantly higher diagnostic reasoning scores than the control group, with an adjusted difference of 7.6 percentage points (95% CI: 1.4 to 13.9; *P*=0.016) from a prespecified linear mixed-effects model.

In a post-hoc analysis stratifying cases by error condition (Supplementary Table 1), the nudge improved diagnostic reasoning accuracy for error-containing recommendations by an adjusted 10.3 percentage points (95% CI, 0.5 to 20.0; *P*=0.039) and for error-free recommendations by 5.0 percentage points (95% CI, -1.5 to 11.6; *P*=0.130), the latter exceeding the prespecified significance threshold (*P*≤0.05).

### Secondary Outcomes

The mean (SD) top-choice diagnosis accuracy was 79.5 (39.1) for the treatment group and 69.1 (45.1) for the control group (Fig. 3, Table 3). The linear mixed-effects model resulted in an adjusted difference of 10.9 percentage points (95% CI, 1.7 to 20.0 pp; *P*=0.020).

**Table 3.**
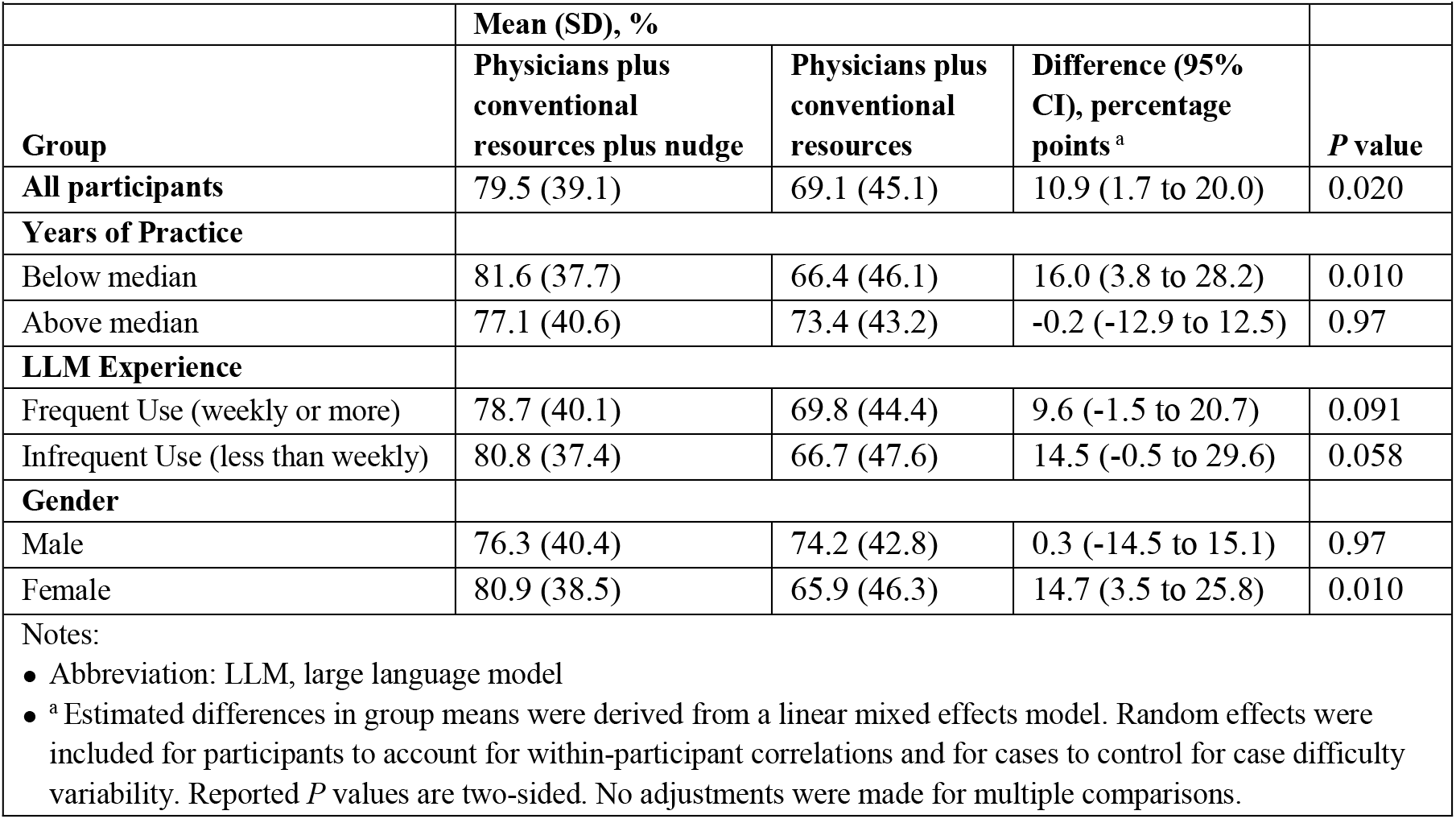
Top-Choice Diagnosis Accuracy Outcome.

| **Table 3. Top-Choice Diagnosis Accuracy Outcome** |  |  |  |  |
| --- | --- | --- | --- | --- |
|  | **Mean (SD), %** |  |  |  |
| **Group** | **Physicians plus conventional resources plus nudge** | **Physicians plus conventional resources** | **Difference (95% CI), percentage points <sup>a</sup>** | ***P* value** |
| **All participants** | 79.5 (39.1) | 69.1 (45.1) | 10.9 (1.7 to 20.0) | 0.020 |
| **Years of Practice** |  |  |  |  |
| Below median | 81.6 (37.7) | 66.4 (46.1) | 16.0 (3.8 to 28.2) | 0.010 |
| Above median | 77.1 (40.6) | 73.4 (43.2) | -0.2 (-12.9 to 12.5) | 0.97 |
| **LLM Experience** |  |  |  |  |
| Frequent Use (weekly or more) | 78.7 (40.1) | 69.8 (44.4) | 9.6 (-1.5 to 20.7) | 0.091 |
| Infrequent Use (less than weekly) | 80.8 (37.4) | 66.7 (47.6) | 14.5 (-0.5 to 29.6) | 0.058 |
| **Gender** |  |  |  |  |
| Male | 76.3 (40.4) | 74.2 (42.8) | 0.3 (-14.5 to 15.1) | 0.97 |
| Female | 80.9 (38.5) | 65.9 (46.3) | 14.7 (3.5 to 25.8) | 0.010 |
| Notes:<br>• Abbreviation: LLM, large language model<br>• <sup>a</sup> Estimated differences in group means were derived from a linear mixed effects model. Random effects were included for participants to account for within-participant correlations and for cases to control for case difficulty variability. Reported *P* values are two-sided. No adjustments were made for multiple comparisons. |  |  |  |  |

**Figure 3:**
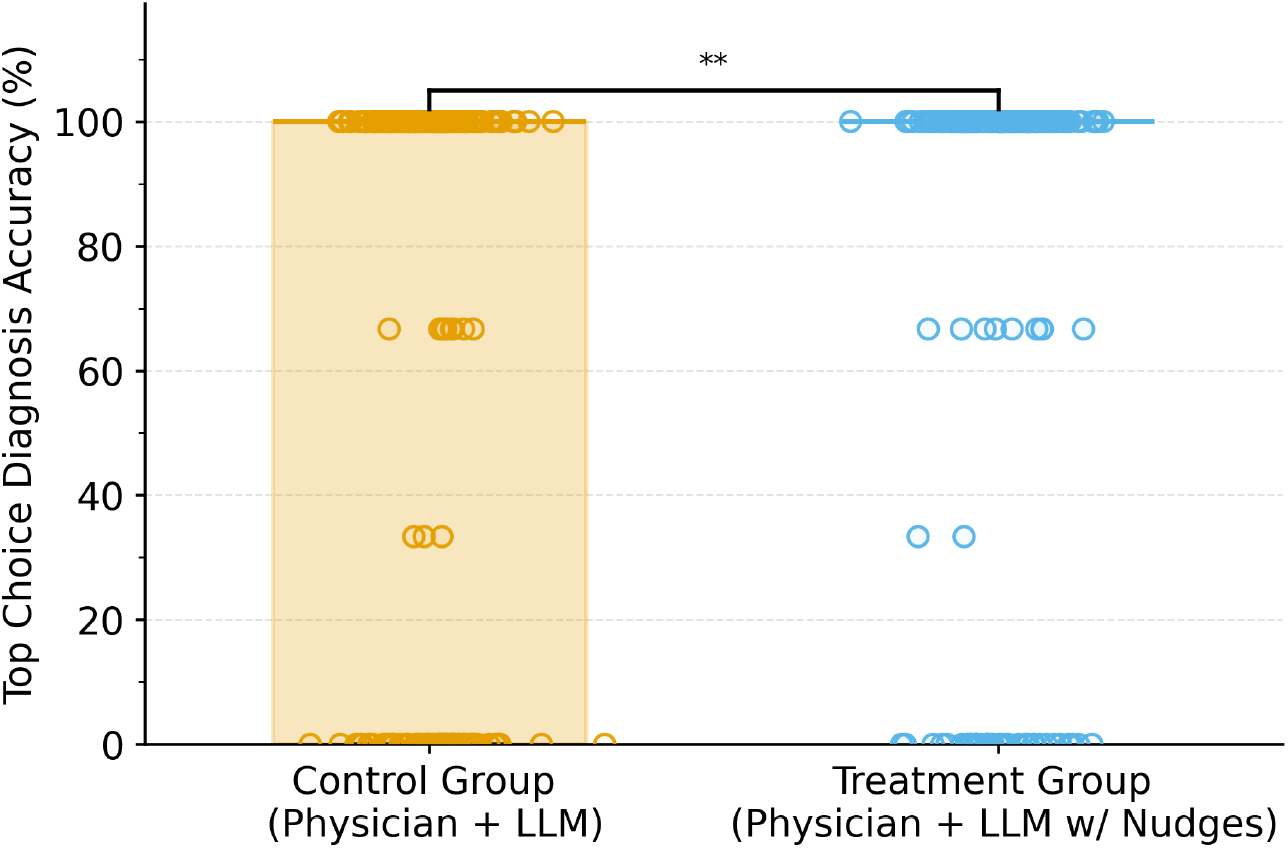
Comparison of the secondary outcome for physicians in the treatment group with physicians in the control group (top-choice diagnosis accuracy score standardized to 0-100). Seventy-two physicians were randomized 1:1 and completed 432 cases (216 in the treatment group, 216 in the control group). The center of the box plot represents the median, with the boundaries representing the first and third quartiles. The whiskers represent the furthest data points from the center within 1.5 times the IQR. Individual data points show case-level scores. The treatment group demonstrated significantly higher average top-choice diagnosis accuracy score than the control group, with an adjusted difference of 10.9 percentage points (95% CI: 1.7 to 20.0; *P*=0.020) from a prespecified linear mixed-effects model.

### Subgroup Analyses

Tables 2 and 3 present treatment-versus-control comparisons stratified by three physician characteristics: (1) years of clinical practice since MBBS, (2) self-reported LLM experience, and (3) gender. We find that the treatment effect was larger among those at or below the median years of practice (8.0 years), who experienced 11.9 percentage points improvement (95% CI, 4.1 to 19.7 pp; *P*=0.0029), compared to a -1.9 percentage points decline among more experienced physicians (95% CI, -10.3 to 6.5 pp; *P*=0.66). Similarly, the treatment effect was greater among frequent users, who showed a 7.0 percentage points increase in diagnostic accuracy (95% CI, -0.3 to 14.3 pp; *P*=0.060), versus a 9.8 percentage point improvement among those reported infrequent users (95% CI, -1.6 to 21.2 pp; *P*=0.092). The treatment benefit also varied significantly by gender, with female physicians experiencing a 12.1 percentage points improvement (95% CI, 4.8 to 19.4 pp; *P*=0.0011) compared to a -3.2 percentage points decrease among male physicians (95% CI -13.3 to 6.8 pp; *P*=0.53). In post-hoc, exploratory analyses that were not prespecified, diagnostic reasoning accuracy stratified by clinical case and by participants’ medical specialty is presented in Supplementary Tables 2 and 3, respectively.

### Assessment Tool Validation

Inter-rater reliability among three graders for the primary outcome was high (Krippendorff’s α = 0.89), consistent with diagnostic performance studies, and internal consistency of the grading instrument was strong (Cronbach’s α = 0.96). Krippendorff’s α and Cronbach’s α for Top-Choice Diagnosis Accuracy is presented in Supplementary Table 4.

## Discussion

In this randomized trial, a lightweight, transparency-based nudge significantly improved diagnostic reasoning among AI-trained physicians consulting LLM recommendations: the dual-component nudge yielded a 7.6 percentage-point gain over the control group, despite identical ChatGPT-5.1 access and AI-literacy preparation. The effect is clinically meaningful given prior evidence that a 20-hour AI-literacy curriculum did not protect physicians from automation bias on recommendations containing subtle, clinically significant errors.^26^ Education alone is therefore insufficient; a scalable point-of-care intervention built around trust calibration can attenuate AI-induced diagnostic error while preserving physician discretion, since LLM consultation remained voluntary. Consistent with this mechanism, the nudge’s benefit was concentrated on recommendations containing embedded errors (Supplementary Table 1), where automation bias poses the greatest patient-safety risk, and was more modest for error-free recommendations.

Several mechanisms may explain why the nudge was effective. The anchoring cue likely addressed a known calibration deficit: physicians’ subjective estimates of LLM accuracy are poorly aligned with empirical performance, and a quantitative benchmark may have shifted their priors toward more accurate baselines.^17-21^ The ensemble confidence signal acted as a selective-attention cue, prompting System 2 engagement precisely where independent models disagreed, the cases where hallucinations are statistically more likely. Selectivity matters because uniform skepticism is cognitively costly and erodes the genuine accuracy gains LLM consultation provides on routine cases.^25^ Unlike previously studied cognitive forcing functions (e.g., commit-first prompts or fixed deliberation delays), which apply friction to every decision and are poorly tolerated,^37,58^ focusing scrutiny only on disagreement preserved low-friction processing of routine recommendations while concentrating effort where it was most needed.^25^ Component-isolation studies should disentangle these contributions.

The ensemble design merits consideration. Unlike conventional AI confidence scores, derived from a single model’s internal probabilities and poorly calibrated for narrative outputs, ensemble disagreement provides a model-agnostic reliability signal independent of any system’s self-assessment. The choice of three distinct families is empirically grounded: in clinical diagnostic benchmarks, multi-agent systems built from dissimilar models outperform single-vendor counterparts, whereas homogeneous teams suffer correlated failure modes.^59^ Agreement among Claude Sonnet 4.5, Gemini 2.5 Pro Thinking, and GPT-4o thus likely reflects well-established knowledge, while disagreement may flag cases involving ambiguous presentations, out-of-distribution reasoning, or domain-specific hallucinations to which a single model is more susceptible. The approach needs no model internals and updates as models improve, though cross-family diversity does not eliminate correlated error,^59^ and multiple queries carry operational cost.

Our findings bear directly on the policy debate over clinical AI safety. Health systems and regulators have largely converged on AI-literacy training as the primary safeguard against automation bias,^27-29^ yet the relevant frictions operate at the point of decision, not the point of training: knowing in the abstract that LLMs can err does not reliably translate into catching a specific plausible-looking error under time pressure. Rather than relying solely on educating clinicians to be skeptical, health systems can build calibration cues directly into the AI-consultation interface, so the disposition instilled by training is triggered when it matters. This parallels human-factors engineering, where interface modifications consistently outperform training-only interventions in high-stakes domains.^60^

Prespecified subgroup analyses suggested the benefit was unevenly distributed: it was concentrated among physicians at or below the median years of practice and not detectable among more experienced physicians, and favored female over male physicians, while benefit was comparable across frequent and infrequent prior LLM users. One interpretation is that more experienced physicians, whose diagnostic reasoning relies more heavily on heuristic (System 1) processing, may be both more susceptible to automation bias and less responsive to calibration cues; ^48-52^ the gender difference lacks a clear mechanistic explanation. Because these comparisons were not powered as primary analyses, whether the nudge protects all physicians equally requires further study.

Time-on-task was suggestive of the hypothesized mechanism: nudged physicians spent longer per case (adjusted difference, 52.5 seconds; 95% CI, -12.1 to 117.1; *P*=0.11), with a larger increment on cases with incorrect LLM recommendations (61.1 seconds; *P*=0.20) than correct ones (43.9 seconds; *P*=0.33; Supplementary Table 5), consistent with selective System 2 engagement rather than uniform slowing from visual salience. However, none of these differences reached statistical significance, and we captured no direct measures of cognitive load; eye-tracking or think-aloud protocols could more definitively adjudicate the cognitive pathway.

Our study has several limitations. Clinical vignettes enable controlled error placement but cannot capture real-world encounters shaped by multimorbidity and incomplete information; prospective deployment studies are needed to confirm generalizability. Rapid LLM development means our findings may evolve, and the single-session design leaves open whether effects persist or physicians habituate to the cues. We also did not evaluate alternative nudge designs, such as commit-first^58^ or forced-delay cognitive forcing functions.^37^ Nonetheless, our results provide the first randomized evidence that automation bias in physician-LLM collaboration is not immutable but a tractable problem responsive to lightweight, scalable intervention. As AI-assisted decision-making becomes embedded in clinical workflows, building trust-calibration cues into the consultation interface may offer a practical path toward preserving the diagnostic judgment that patient safety depends on.

## Methods

Our study was approved by the Lahore University of Management Sciences (LUMS) Institutional Review Board, and all participants provided informed consent prior to enrollment. Participants received PKR 5,000 (approximately USD 20) as compensation for participating in the study. The trial was prospectively registered on ClinicalTrials.gov (NCT07328815; first submitted December 26, 2025 and first posted January 9, 2026) before participant enrollment. The study design and reporting adhere to the Consolidated Standards of Reporting Trials (CONSORT) 2025 guidelines; the complete study protocol is in the supplementary information (Supplementary Note 1).

Physicians with varying specialties and clinical experience were recruited through email distribution lists at the LUMS Learning Institute, which offers specialized training programs for physicians on health care AI and data science. Participants represented 47 medical institutions across Pakistan. Eligible participants included physicians registered with the Pakistan Medical and Dental Council who held a Bachelor of Medicine, Bachelor of Surgery (MBBS) degree and had completed a 20-hour AI-literacy training (Supplementary Table 6) covering LLM capabilities, prompt engineering, and strategies for critically evaluating AI-generated output. Participants were recruited from cohorts of the AI literacy training program and were supervised by study coordinators in either remote sessions or at an in-person computer laboratory at LUMS. Each session lasted 85 minutes, comprising a 10-minute baseline survey followed by 75 minutes of clinical vignette assessments.

Participants were randomly assigned (1:1) to either the treatment group (n=36) or the control group (n=36) using a computer-generated randomization sequence created with the Sealed Envelope program, as prespecified in the analysis plan (Supplementary Note 1). To prevent priming effects and ensure valid measurement of genuine, unconscious automation bias, participants were blinded to the study’s specific aims. The single-blind randomization was known only to the study administrator. Fig. 1 illustrates the participant flow, with a visual representation provided in Supplementary Fig. 1.

### Clinical Vignettes

Three physician co-authors (M.A.K, M.J.A., and A.Z.S.) initially developed seven clinical vignettes spanning internal medicine, family medicine, endocrinology and emergency medicine. From this pool, six were chosen that offered a meaningful diagnostic challenge, excluding cases that were overly simple or rare. To ensure consistency and uniform presentation of information, all LLM-generated recommendations were pre-generated and standardized. Pilot testing established a 75-minute study time limit.

The three physician co-authors embedded subtle but clinically significant errors into the ChatGPT-5.1 recommendations for three of the six vignettes, leaving the recommendations for the other three error-free. These errors were designed to be detectable by competent physicians but not immediately apparent on casual review. Both groups evaluated the same six vignettes with the same set of recommendations, three flawed and three accurate, and the vignettes containing flawed recommendations were assigned at random to mitigate pattern recognition. Each vignette followed a standardized format (chief complaint, history of present illness, relevant past medical history, physical examination findings, and laboratory results) to present information uniformly. A sample case is available in Supplementary Table 7.

While we measured top-choice diagnosis accuracy score as a secondary outcome, our primary endpoint was the diagnostic reasoning accuracy, which measured the quality of the diagnostic reasoning process. This was evaluated using structured reflection methodology that mirrored clinical practice. Using a standardized template (Supplementary Table 7), participants documented their top three differential diagnoses with supporting and opposing evidence, top-choice diagnosis with justification, and recommended next steps.^34^ This comprehensive assessment of the reasoning pathway, rather than just the final answer, aligns with recent literature.^11,12^

### Assessing Diagnostic Performance

Participants’ assessment grids were scored by three physicians using a detailed rubric (Supplementary Table 8 and Supplementary Table 9). To ensure objectivity, evaluators were blinded to group assignments, and all identifying metadata was removed from the responses. The rubric comprised five components. The quality of the differential diagnosis was scored from 0 to 3 points, reflecting the number of reasonable differential diagnoses listed and whether the correct diagnosis was included. For each of the three differential diagnoses, the supporting and opposing findings were each awarded 1 point when correctly identified (up to 3 points for supporting findings and 3 points for opposing findings). The top-choice diagnosis was awarded 18 points when correct and 9 points when partially correct, while an incorrect diagnosis received no points. Finally, the appropriateness of the recommended next steps was scored from 0 to 1 point per step (0 for inappropriate or unnecessary steps, 0.5 for partially appropriate steps, and 1 for appropriate and comprehensive steps).

### Study Design

We employed a randomized, single-blind, two-arm parallel-group design. Participants were randomized 1:1 to a treatment or control arm and evaluated six clinical vignettes within a 75-minute window. Both arms used the same custom web platform, which displayed each vignette alongside pre-generated ChatGPT-5.1 diagnostic recommendations; three of the six recommendations contained deliberately introduced, clinically significant errors, and vignettes were presented in randomized order to prevent pattern detection. A key feature of the design was physician autonomy: consulting the AI for any vignette was a voluntary, opt-in action requiring an explicit click to view the output (Supplementary Figs. 2 and 3).

The two arms differed only in the context shown alongside the recommendations. The treatment arm received a dual-component behavioral nudge embedded in the recommendation interface: (1) an anchoring cue displaying ChatGPT’s diagnostic accuracy on standard medical benchmarks, presented before each recommendation to calibrate baseline expectations; and (2) a selective-attention cue in the form of a case-specific signal that displays the mean of three confidence ratings in the accuracy of ChatGPT-5.1 recommendations independently assigned by three LLMs from distinct model families (Claude Sonnet 4.5, Gemini 2.5 Pro Thinking, and GPT-4o) using a standardized prompt (Supplementary Table 10), together with a color-coded traffic-light signal derived from the mean confidence score. A red signal indicated mean ensemble confidence below ChatGPT’s benchmark accuracy, flagging a high-uncertainty recommendation that warranted closer scrutiny; an orange signal indicated above-baseline but sub-maximal confidence; and a green signal indicated full ensemble consensus, accompanied by a standard caution that AI may make mistakes (Supplementary Figs. 4 and 5). The confidence score displayed for each ChatGPT-5.1 recommendation is reported in Supplementary Table 11; per-model confidence scores from the three-model ensemble under the correct-version and error-induced conditions are provided in Supplementary Tables 12 and 13, respectively. The control arm viewed the ChatGPT-5.1 recommendations without the nudge (Supplementary Fig. 6).

Both groups had access to conventional online medical resources, such as medical databases and standard search, to support their diagnostic workflow; however, to isolate the intervention’s effect and prevent confounding AI exposure, a browser extension specifically blocked Google’s “AI Overviews.” Participants were instructed to approach each clinical vignette as they would in their regular clinical practice, and all diagnostic assessments were collected electronically via a secure custom web platform. No concomitant care was applicable, as participants were physicians completing assessments, not patients. Because the intervention was judged a priori to pose no more than minimal risk, we did not pre-specify individual adverse-event categories; harms were assessed non-systematically by recording withdrawals and incidents during test sessions.

### Validation of Assessment Tool

To finalize the assessment rubrics, three licensed physicians with expertise in clinical reasoning assessment (M.A.K, M.J.A., and A.Z.S.) independently solved each clinical vignette to flag potential scoring discrepancies. Disagreements were resolved through structured consensus discussions, resulting in standardized scoring rubrics for each case that accounted for clinical ambiguity by allowing for multiple correct variations if supported by expert consensus. Subsequently, each participant’s responses underwent blinded evaluation by three physicians. Inter-rater reliability was assessed using Krippendorff’s alpha, and the instrument’s internal consistency was measured with Cronbach’s alpha (Supplementary Table 4).

### Study Outcomes

Our prespecified primary outcome was the diagnostic reasoning accuracy, calculated as a percentage of total points achieved on the assessment tool. To determine this score, three blinded physicians (M.A.K., M.J.A., and A.Z.S.) independently scored each response using a pre-defined rubric they developed. The final score for each case was the arithmetic mean of the three scores. The prespecified secondary outcome was the top-choice diagnosis accuracy, the correctness of the physician’s single most likely diagnosis for each vignette. All outcomes were compared between the randomized groups at the case-level.

### Statistical Analysis

Demographic and baseline characteristics were compared between groups using χ^2^ or Fisher exact tests for categorical variables and two-sided *t*-test or Mann-Whitney *U* test for the mean and median of continuous variables, respectively.

Our target sample size was 50 participants (25 per arm), based on a prior study.^11^ An a priori power analysis, conducted using Python version 3.11.9 and statsmodels version 0.14.4, indicated that 200 completed cases (4 per participant) would provide at least 80% power to detect an 8-percentage-point mean difference in scores, assuming a two-sided α of 0.05. The analysis employed mixed-effects models suitable for cluster-randomized designs, considering an intraclass correlation coefficient ranging from 0.05 to 0.15 and standard deviation of 16.2%. Ultimately, 72 participants enrolled and completed the study, yielding 432 completed cases (6 per participant).

All analyses followed the intention-to-treat principle and were conducted at the case level, with cases clustered by participants. We used linear mixed-effects models to evaluate differences in primary and secondary outcomes, with adjustment for baseline covariates (years of practice post-MBBS, prior LLM experience, and gender). Random effects were included for participants to account for within-participant correlations and for cases to control for case difficulty variability (Supplementary Note 2). To assess the sensitivity of our findings to model specification and distributional assumptions, we compared our primary mixed-effects model results against ordinary least squares (OLS) with clustered standard errors, non-parametric cluster bootstrap resampling at the physician level, and models without baseline covariate adjustment. Results were consistent across specifications for both the primary and secondary outcomes. These sensitivity analyses and robustness checks are available in Supplementary Tables 14, 15 and 16, and Supplementary Fig. 7. As no multiplicity adjustment was prespecified for secondary endpoints or subgroup comparisons (by years of practice, LLM experience, and gender), these are reported descriptively as mean differences with 95% confidence intervals. In accordance with the statistical analysis plan, these results should not be interpreted as formal hypothesis tests. Stratification by clinical case and by participants’ medical specialty was post-hoc and exploratory, undertaken after data collection, and is reported descriptively without inferential claims.

All statistical analyses were conducted using Python (version 3.11.12) with the pandas library for data manipulation and statsmodels (version 0.14.4) for mixed-effects modeling. The prespecified statistical analysis plan was uploaded to ClinicalTrials.gov (NCT07328815; December 26, 2025) and is also provided in Supplementary Note 1.

### Inclusion & Ethics

All authors meet Nature Portfolio authorship criteria. Prior to the study, Institutional Review Board approval was obtained, and the study was prospectively registered at ClinicalTrials.gov (NCT07328815). A completed CONSORT 2025 checklist accompanies the submission. Throughout, we followed institutional research-integrity guidelines, the Global Code of Conduct for Research in Resource-Poor Settings, and inclusive citation practices to ensure that no groups are disadvantaged.

## Data Availability

De-identified participant data that support the findings of this study will be made publicly available in an open repository upon publication of this article.

## Acknowledgement

This work was supported by the Learning Institute at Lahore University of Management Sciences (I.A.Q. and M.H.A.). We thank Amna Hassan and Laiba Intizar Ahmad for administering the surveys and proctoring study sessions, who served as a paid research assistant on this project and received compensation from study funds.

## Author Contributions

I.A.Q., A.A., and M.H.A. contributed to concept and design. I.A.Q., A.A., A.U.K., A.Z.S., M.J.A., and M.H.A. contributed to acquisition, analysis, or interpretation of data. I.A.Q. and A.A. contributed to drafting of the manuscript. I.A.Q. and A.A. contributed to statistical analysis. I.A.Q. and M.H.A. contributed to obtaining funding. I.A.Q., A.A., A.U.K., A.Z.S., M.J.A., and M.H.A. contributed to administrative, technical, or material support. I.A.Q., A.A., and M.H.A. contributed to supervision. All authors reviewed the manuscript.

## Patient and Public Involvement

Not applicable. This study involved only physicians as participants and aimed to evaluate clinical decision-making performance using standardized vignettes. No patients were recruited as research participants.

