## Supplementary Information for "Mitigating Automation Bias in Physician-LLM Diagnostic Reasoning Using Behavioral Nudges: A Randomized Controlled Trial"

A Randomized Clinical Trial

Ihsan Ayyub Qazi, PhD; Ayesha Ali, PhD; Asad Ullah Khawaja, MBBS; Muhammad Junaid Akhtar, MBBS; Ali Zafar Sheikh, MBBS; Muhammad Hamad Alizai, PhD

This supplementary material has been provided by the authors to give readers additional

information about their work.

### Supplementary Table 1: Treatment Effect by LLM Recommendation Accuracy

| **Diagnostic Reasoning Accuracy (%)** | | | | |
| --- | --- | --- | --- | --- |
|  | **Raw Mean, %** | |  |  |
| **Condition** | **Treatment**  (Physicians plus conventional resources plus nudge | **Control**  (Physicians plus conventional resources) | **Difference (95% CI), percentage points** ^a^ | ***P* value** |
| **No-Error LLM Recommendation** | 90.6 | 85.9 | 4.8 (−0.5 to 10.1) | 0.077 |
| **Error LLM Recommendation** | 75.0 | 65.3 | 9.7 (1.7 to 17.7) | 0.017 |
|  | **Model Estimated Mean, %** | | **Difference (95% CI), percentage points** ^b^ |  |
| **No-Error LLM Recommendation** | 92.3% | 87.3% | 5.0 (−1.5 to 11.6) | 0.130 |
| **Error LLM Recommendation** | 76.7% | 66.4% | 10.3 (0.5 to 20.0) | 0.039 |
| Notes:   - Abbreviations: LLM, large language model - Raw means are observed (raw) group means of the total score (%); each group n = 108 (216 cases per condition). - For model estimated means, separate linear mixed-effects models were fitted for cases with no-error (correct) and error (incorrect) LLM recommendations. - ^a^ The ‘Difference’ (top panel) is the unadjusted difference in means between the Treatment and Control groups for each condition; 95% CI and two-sided *P* value are from a two-sample test allowing unequal variances (Welch). No adjustment was made for multiple comparisons. - ^b^ The ‘Difference’ is the adjusted (Treatment - Control) effect from a separate linear mixed-effects model for each condition, adjusting for gender, years of practice, and LLM experience, with random intercepts for participant and case; it therefore need not equal the simple difference of the displayed means (unadjusted differences: 4.78 [no error] and 9.69 [error]). Reported *P* values are two-sided; no adjustment was made for multiple comparisons. | | | | |

### Supplementary Table 2: Diagnostic Reasoning Accuracy Stratified by Case

| **Case #** | **Treatment**  (Physicians plus conventional resources plus nudge | **Control**  (Physicians plus conventional resources) | **Difference (95% CI), percentage points** ^a^ | **N**  (Treatment) | **N**  (Control) |
| --- | --- | --- | --- | --- | --- |
| 1 | 90.4 (15.4) | 81.1 (25.5) | 9.4 (-0.6 to 19.3) | 36 | 36 |
| 2 | 67.8 (30.3) | 69.4 (31.8) | -1.6 (-16.2 to 13.0) | 36 | 36 |
| 3 | 81.7 (28.6) | 73.0 (32.1) | 8.7 (-5.6 to 23.0) | 36 | 36 |
| 4 | 89.6 (16.8) | 80.4 (22.5) | 9.2 (-0.2 to 18.5) | 36 | 36 |
| 5 | 83.1 (21.2) | 81.4 (22.3) | 1.7 (-8.5 to 11.9) | 36 | 36 |
| 6 | 84.1 (26.8) | 68.1 (34.0) | 16.0 (1.6 to 30.4) | 36 | 36 |
| Notes:   - Abbreviation: LLM, large language model - ^a^ Post-hoc exploratory analysis; not prespecified. Unadjusted mean differences in diagnostic reasoning accuracy for each case, with 95% confidence intervals calculated using two-sided t-tests. The widths of these intervals have not been adjusted for multiplicity and should not be interpreted as hypothesis tests. | | | | | |

### Supplementary Table 3: Diagnostic Reasoning Accuracy by Medical Specialty

| **Physician Category** | **Treatment**  (Physicians plus conventional resources plus nudge | **Control**  (Physicians plus conventional resources) | **Difference (95% CI), percentage points** ^a^ | **N** (Treatment) | **N** (Control) |
| --- | --- | --- | --- | --- | --- |
| Physician (primary care & medical specialties) | 86.2 (20.8) | 77.9 (26.1) | 8.3 (1.9 to 14.6) | 108 | 108 |
| Surgeon | 79.2 (28.6) | 69.6 (33.7) | 9.6 (-1.6 to 20.7) | 78 | 54 |
| Diagnostic/support specialty | 87.4 (27.1) | 91.2 (21.5) | -3.8 (-32.9 to 25.3) | 6 | 12 |
| Academic / Non-Clinical | 78.1 (26.4) | 72.7 (28.2) | 5.4 (-8.5 to 19.3) | 24 | 42 |
| Notes:   - Abbreviation: LLM, large language model - ^a^ Post-hoc exploratory analysis; not prespecified. Unadjusted mean differences in diagnostic reasoning accuracy for each medical specialty category, with 95% confidence intervals calculated using two-sided t-tests. The widths of these intervals have not been adjusted for multiplicity and should not be interpreted as hypothesis tests. | | | | | |

### Supplementary Table 4. Inter-Rater Reliability & Internal Consistency for Diagnostic Reasoning Accuracy & Top-Choice Diagnosis Accuracy

| **Component** | **Krippendorff's Alpha** | **Cronbach’s Alpha** |
| --- | --- | --- |
| Top-Choice Diagnosis Accuracy  (*Secondary Outcome*) | 0.911 | 0.969 |
| Diagnostic Reasoning Accuracy  (*Primary Outcome*) | 0.889 | 0.961 |

*Caption: Inter-rater reliability and internal consistency for Top-Choice Diagnosis Accuracy and Diagnostic Reasoning Accuracy. Krippendorff's α quantifies inter-rater agreement and Cronbach's α quantifies internal consistency across raters for each scoring component. Each of the 432 case-level units (72 participants × 6 clinical cases) was independently scored by three raters. Cronbach's α treats raters as items; Krippendorff's α is computed over the rater-by-unit matrix. For Cronbach's α, values ≥ 0.70 indicate acceptable internal consistency. For Krippendorff's α, values ≥ 0.80 indicate reliable agreement; both components here exceed 0.80, indicating strong inter-rater agreement.*

### Supplementary Table 5: Time Spent Per Case

| **Time Spent Per Case Outcome (seconds)** | | | | |
| --- | --- | --- | --- | --- |
|  | **Mean, sec** | |  |  |
| **Condition** | **Treatment**  (Physicians plus conventional resources plus nudge | **Control**  (Physicians plus conventional resources) | **Difference (95% CI), seconds^a^** | **P value** |
| **Overall** | 631.1 | 578.6 | 52.5 (-12.1 to 117.1) | 0.111 |
| **Incorrect LLM recommendations** | 675.3 | 614.2 | 61.1 (-32.5 to 154.7) | 0.201 |
| **Correct LLM recommendations** | 586.9 | 543.0 | 43.9 (-43.6 to 131.4) | 0.325 |
| Notes:   - Abbreviation: LLM, large language model. - Treatment group received nudges; Control group did not. The ‘Difference’ is the estimated difference in mean time spent per case (Treatment − Control), in seconds. - ^a^The overall estimate for ‘Difference’ was derived from a linear mixed effects model adjusting for gender, years of practice, and LLM experience, with random intercepts for participant and case. For the incorrect-recommendation and correct-recommendation conditions, separate mixed effects models were fitted. Reported *P* values are two-sided. No adjustments were made for multiple comparisons. | | | | |

### Supplementary Table 6. AI Training Topics.

| **AI Training Topics** | |
| --- | --- |
| 1. | **Introduction to Artificial Intelligence (AI)**   - What is AI and why it matters - AI as a helpful assistant: from mechanical tasks to intelligent support |
| 2. | **Basics of Machine Learning (ML)**   - What is machine learning and how it works - Overview of ML types - Real-life examples, including healthcare applications |
| 3. | **Basics of Generative AI (GenAI) and Prompt Engineering**   - How GenAI works in simple terms - Introduction to prompt engineering techniques |
| 4. | **Writing and Research using GenAI**   - Writing professional and effective emails with AI assistance - Retrieval-Augmented Generation (RAG) for improved outputs |
| 5. | **AI-powered Presentation Design**   - Creating presentations from descriptions and documents - Prompt engineering techniques specific to presentation generation |
| 6. | **Multimedia Content Creation with GenAI**   - Generating and editing images using AI - Creating audio and video content using simple prompts - Digitizing and summarizing medical notes with AI |
| 7. | **GenAI for Data and Math Tasks**   - Using GenAI to explore and understand data - Prompting for mathematical and data analysis tasks |
| 8. | **GenAI Customization and Limitations**   - Using and building your own custom GPTs - Understanding AI limitations like hallucinations and how to manage them |

### Supplementary Table 7. Diagnostic Case-6: Vignette and Questions

**Diagnostic Vignette**

**History of Present Illness**

A 52-year-old male presents with a 3-week history of worsening fatigue, low-grade fevers, non-productive cough that progressed to streaks of blood in the sputum over the past 4 days, and bilateral knee and ankle arthralgias. He reports persistent nasal congestion with occasional epistaxis for several months. He denies recent travel, sick contacts, or new medications. He has unintentionally lost 4 kg in the last month.

**Past Medical History**

Hypertension (well controlled on amlodipine). Seasonal allergic rhinitis. Former smoker (20 pack-years; quit 5 years ago). No known kidney disease; serum creatinine was 0.9 mg/dL at an annual check-up 8 weeks ago. Family history non-contributory.

**Physical Examination**

VITALS: BP 148/92 mmHg, HR 98 bpm, RR 20/min, Temp 37.6 °C (99.7 °F), SpO2 93% on room air.

GEN: Appears fatigued, mild respiratory distress with conversation.

ENT: Crusted nasal mucosa with mild septal tenderness; no oral ulcers.

CARDIAC: Regular rhythm, no murmurs.

PULM: Bibasilar crackles; no wheezes.

ABD: Soft, non-tender.

EXT: No edema, mild tenderness over knees and ankles without effusion.

SKIN: No rash, purpura, or ulcers.

**Laboratory Results**

Hemoglobin 9.8 g/dL (baseline 13.5); WBC 12.1 × 103/μL; Platelets 310 × 103/μL; ESR 88 mm/hr;

CRP 68 mg/L.

Serum creatinine 3.9 mg/dL; BUN 64 mg/dL; potassium 5.2 mmol/L; bicarbonate 18 mmol/L. Urinalysis: 3+ blood, 1+ protein; microscopy: numerous dysmorphic RBCs, red cell casts.

Serologies: c-ANCA positive (PR3 112 U/mL); p-ANCA negative; anti-GBM antibody <3 U/mL (normal < 20); ANA negative; C3/C4 normal.

Chest X-ray: Multiple bilateral cavitary nodules and patchy alveolar opacities.

**Structured Diagnostic Reasoning Grid**

**PART 1: Differential Diagnosis**
**Please provide three possible diagnoses with supporting and opposing evidence for each**

**A. Diagnosis 1:**

a. Supporting Evidence for Diagnosis 1:

b. Opposing Evidence for Diagnosis 1:
 .

**B. Diagnosis 2:**

a. Supporting Evidence for Diagnosis 2:

b. Opposing Evidence for Diagnosis 2:

**C. Diagnosis 3:**

a. Supporting Evidence for Diagnosis 3:

b. Opposing Evidence for Diagnosis 3:

**PART 2 - Final Diagnosis (or Top Choice Diagnosis)**

Based on your assessment, select your most likely diagnosis and provide its justification.

a. Final Diagnosis:

b. Justification for Final Diagnosis:

**PART 3 - Additional Steps:**

**List up to three additional steps you would take in the diagnostic or management plan for this patient.**

Step 1

Step 2

Step 3

### Supplementary Table 8. Structured Reflection Rubric for Diagnostic Case-6 – High-Scoring Example Response

| **Question** | **High Scoring Example** | **Score** |
| --- | --- | --- |
| **Question 1:**  **Differential Diagnosis**  List 3 Possible Diagnoses  (1 point each) | 1. Granulomatosis with polyangiitis (GPA, Wegener’s)  2. Microscopic polyangiitis (MPA)  3. Anti-GBM antibody disease (Goodpasture syndrome) | 3/3 |
| **Question 2:**  **Supporting Evidence**  For each possible diagnosis, provide findings/risk factors supporting this hypothesis  (1 point each) | **1. GPA:** a) c-ANCA/PR3 strongly positive, b) Upper and lower respiratory tract involvement (chronic sinus symptoms, cavitary pulmonary nodules, hemoptysis) and c) Rapidly progressive glomerulonephritis with RBC casts.  **2. MPA:** a) Pulmonary-renal syndrome (hemoptysis + RPGN), b) Pauci-immune ANCA-associated pattern (even though p-ANCA negative, MPA can be ANCA-positive or negative) and c) Constitutional symptoms and arthralgias.  **3. Anti-GBM antibody disease:** a) Pulmonary-renal syndrome with hematuria and hemoptysis and b) Rapid rise in creatinine from 0.9 mg/dL to 3.9 mg/dL within 2 months. | 3/3 |
| **Question 3**  **Opposing Evidence**  For each possible diagnosis, provide findings opposing this hypothesis, or findings that were expected but not present  (1 point each) | **1. GPA:** a) No biopsy yet confirming granulomatous inflammation and b) Lack of skin purpura or ENT ulceration typical of severe GPA.  **2. MPA:** a) Negative p-ANCA/MPO serology and b) Presence of cavitary nodules favors GPA over MPA.  **3. Anti-GBM antibody disease:** a) Anti-GBM antibody negative and b) Cavitary lung lesions are atypical for Goodpasture. | 3/3 |
| **Question 4:**  **Final Diagnosis**  (18 points) | **Granulomatosis with polyangiitis (Wegener’s).**  c-ANCA/PR3 strongly positive, upper and lower respiratory tract involvement (chronic sinus symptoms, cavitary pulmonary nodules, hemoptysis), rapidly progressive glomerulonephritis with RBC casts. | 18/18 |
| **Question 5:**  **Additional Steps**  (1 point each) | • Urgent kidney biopsy to confirm pauci-immune necrotizing crescentic glomerulonephritis.  • Initiate high-dose IV methylprednisolone pulses followed by oral prednisone taper.  • Consider plasma exchange if severe alveolar hemorrhage or rapidly worsening renal failure. | 3/3 |
| **Total Score** |  | 30/30 |

*Caption: Example of a high-scoring response. The participant provided three plausible diagnoses with accurate and comprehensive supporting and opposing evidence and reached the correct final diagnosis.*

### Supplementary Table 9. Structured Reflection Rubric for Diagnostic Case-6 – LLM Recommendation with Deliberate Errors (Low-Scoring Example Response)

| **Question** | **Low Scoring Example** | **Score** |
| --- | --- | --- |
| **Question 1:**  **Differential Diagnosis**  List 3 Possible Diagnoses  (1 point each) | 1. Poly Arteritis Nodosa (PAN)  2. Granulomatosis with polyangiitis (GPA, Wegener’s)  3. Anti-GBM antibody disease (Goodpasture syndrome) | 2/3 |
| **Question 2:**  **Supporting Evidence**  For each possible diagnosis, provide findings/risk factors supporting this hypothesis  (1 point each) | **1. PAN:** a) Overwhelming renal involvement and b) joint involvement  **2. GPA:** a) c-ANCA/PR3 strongly positive. b) Upper and lower respiratory tract involvement with cavitary nodules and hemoptysis.  **3. Anti-GBM antibody disease:** a) Pulmonary-renal syndrome with hematuria and hemoptysis. b) Rapid rise in creatinine. | 2/3 |
| **Question 3**  **Opposing Evidence**  For each possible diagnosis, provide findings opposing this hypothesis, or findings that were expected but not present (1 point each) | **1. PAN:** a) Positive c-ANCA b) cavitatory nodules on chest X-ray  **2. GPA:** a) No biopsy yet confirming granulomatous inflammation.  **3. Anti-GBM antibody disease:** a) Anti-GBM antibody negative. b) Cavitary lung lesions are atypical. | 3/3 |
| **Question 4:**  **Final Diagnosis**  (18 points) | **Poly Arteritis Nodosa (PAN)**. Based on renal involvement and joint involvement. | 0/18 |
| **Question 5:**  **Additional Steps**  (1 point each) | • Urgent kidney biopsy.  • high-dose IV methylprednisolone pulses followed by oral prednisone taper.  • Consider plasma exchange if severe alveolar hemorrhage or rapidly worsening renal failure. | 3/3 |
| **Total Score** |  | 10/30 |

*Caption: Example of a LLM recommendation containing deliberate errors, resulting in a low-scoring response. The grading reflects the score assigned by a one of the three physicians in the panel.*

### Supplementary Table 10: Prompt for Generating Confidence Scores by Three LLMs

**Prompt**

Role: You are an experienced board-certified physician in the relevant specialty.

Task: You will be shown (1) a detailed clinical vignette and (2) a physician’s diagnostic reasoning or management recommendation. Your job is to rate how accurate and appropriate the physician’s response is, given the clinical details provided.

Instructions:

1. Read the vignette carefully to identify key clinical features, lab results, and imaging findings.

2. Assess whether the provided response:

- Correctly identifies the most likely diagnosis based on evidence.

- Demonstrates sound clinical reasoning (uses appropriate supporting and opposing findings).

- Recommends appropriate next steps for management or investigation.

3. Assign a confidence score from 1 to 100, where:

90-100 → Strongly confident; highly accurate and well-reasoned.

70-89 → Generally correct but minor gaps or less complete reasoning.

50-69 → Partially correct; some reasoning errors or missed key findings.

30-49 → Major diagnostic or management errors.

1-29 → Fundamentally incorrect or unsafe clinical reasoning.

Output Format:

- Confidence Score: [number between 1-100].

- Brief Justification (1-3 sentences): Explain which aspects of the response were accurate or flawed.

Vignette: …

### Supplementary Table 11: Confidence Scores of LLM (ChatGPT-5.1) Recommendations

Mean confidence score (%) of Claude 4.5, Gemini 2.5, and GPT-4o, by clinical scenario (ID 1-6).

| **ID** | **Correct version** | **Error-induced** | **Δ (Correct - Error)** |
| --- | --- | --- | --- |
| 1 | 98.3 | 50.7 | 47.6 |
| 2 | 95.0 | 60.7 | 34.3 |
| 3 | 97.3 | 35.0 | 62.3 |
| 4 | 95.7 | 35.0 | 60.7 |
| 5 | 94.0 | 57.3 | 36.7 |
| 6 | 93.3 | 41.7 | 51.6 |
| Notes: Values are mean confidence scores (%) averaged across the three evaluated models (Claude 4.5, Gemini 2.5, and GPT-4o) for each scenario. The “Correct version” column reflects confidence in LLM recommendations generated by GPT-5.1, whereas the “Error-induced” column reflects confidence in LLM recommendations after deliberate errors (or inaccuracies) were introduced by the panel of physicians. The Δ column denotes the absolute difference between the two conditions (percentage points). Higher Δ values indicate greater erosion of model confidence under error-induced conditions. Per-model values are provided in Tables S14 and S15. | | | |

### Supplementary Table 12: Per-model confidence scores under the correct-version condition.

Confidence score (%) of each evaluated model for recommendations generated from accurate clinical inputs, by clinical scenario (case ID 1-6).

| **Case ID** | **GPT-4o** | **Claude 4.5** | **Gemini 2.5** |
| --- | --- | --- | --- |
| 1 | 100 | 95 | 100 |
| 2 | 90 | 95 | 100 |
| 3 | 97 | 95 | 100 |
| 4 | 95 | 92 | 100 |
| 5 | 90 | 92 | 100 |
| 6 | 90 | 92 | 98 |
| Notes: Values are individual model confidence scores (%) for each scenario under the correct-version condition (GPT-5.1 recommendations for the case without modifications). Row means across the three models correspond to the “Correct version” column of Table S13. | | | |

### Supplementary Table 13: Per-model confidence scores under the error-induced condition.

Confidence score (%) of each evaluated model for recommendations after deliberate errors were introduced, by clinical scenario (case ID 1-6).

| **Case ID** | **GPT-4o** | **Claude 4.5** | **Gemini 2.5** |
| --- | --- | --- | --- |
| 1 | 70 | 42 | 40 |
| 2 | 80 | 42 | 60 |
| 3 | 35 | 35 | 35 |
| 4 | 30 | 35 | 40 |
| 5 | 90 | 42 | 40 |
| 6 | 50 | 35 | 40 |
| Notes: Values are individual model confidence scores (%) for each scenario under the error-induced condition, in which deliberate errors were introduced into the GPT-5.1 recommendations for the case. Row means across the three models correspond to the “Error-induced” column of Table S13. | | | |

### Supplementary Table 14. Sensitivity Analysis of Treatment Effects Across Model Specifications

| **Model** | **Coefficient** | **Standard Error** | **95% CI Lower** | **95% CI Upper** | ***P*-value** | **SE vs Original** |
| --- | --- | --- | --- | --- | --- | --- |
| Mixed Effects (Original) | 7.64 | 3.30 | 1.17 | 14.12 | 0.021 | 1.00 |
| Mixed Effects (Bootstrap) | 7.71 | 3.48 | 0.76 | 14.51 | 0.017 | 1.05 |
| OLS Clustered SE | 7.96 | 3.27 | 1.55 | 14.37 | 0.015 | 0.99 |

*Caption: Treatment effect estimates from mixed-effects regression, bootstrap resampling, and ordinary least squares with clustered standard errors (432 completed cases, 72 participants). The coefficient represents percentage point change in diagnostic accuracy scores. Bootstrap estimates based on 1000 iterations with participant-level resampling. Standard errors are expressed relative to the original mixed-effects model. All approaches show consistent evidence of significant positive treatment effects ranging from 7.64 to 7.96 percentage points, with increasing conservatism in uncertainty estimates across methods. P-values test the null hypothesis of no treatment effect using normal approximation (mixed-effects, OLS) or bootstrap distribution. Sensitivity analysis confirms the robustness of our findings to model specification and distributional assumptions.*

### Supplementary Table 15: Diagnostic Reasoning Accuracy Without Baseline Controls

|  | **Mean (SD), %** | | |  |
| --- | --- | --- | --- | --- |
| **Group** | **Physicians plus conventional resources plus nudge** | **Physicians plus conventional resources** | **Difference (95% CI), percentage points** ^a^ | ***P* value** |
| **All participants** | 82.8 (24.7) | 75.6 (28.7) | 7.2 (1.1 to 13.4) | 0.0207 |
| **Years of Practice** |  | | | |
| Below median | 83.9 (23.1) | 73.0 (28.7) | 10.9 (2.8 to 19.0) | 0.0086 |
| Above median | 81.6 (26.5) | 79.6 (28.4) | 2.0 (-7.1 to 11.0) | 0.6698 |
| **LLM Experience** |  | | | |
| Frequent Use (weekly or more) | 82.4 (22.9) | 73.0 (31.4) | 9.4 (-2.1 to 21.0) | 0.1091 |
| Infrequent Use (less than weekly) | 83.0 (25.8) | 76.3 (27.9) | 6.7 (-0.6 to 14.1) | 0.0728 |
| **Gender** |  | | |  |
| Male | 77.5 (26.8) | 79.5 (27.5) | -2.0 (-12.9 to 8.9) | 0.7224 |
| Female | 85.1 (23.5) | 73.1 (29.2) | 12.1 (5.0 to 19.1) | 0.0008 |
| Notes:   - Abbreviation: LLM, large language model - ^a^ Estimated differences in group means were derived from a linear mixed effects model. Random effects were included for participants to account for within-participant correlations and for cases to control for case difficulty variability. Reported *P* values are two-sided. No adjustments were made for multiple comparisons. | | | | |

### Supplementary Table 16: Top-Choice Diagnosis Accuracy Without Baseline Controls

|  | **Mean (SD), %** | | |  |
| --- | --- | --- | --- | --- |
| **Group** | **Physicians plus conventional resources plus nudge** | **Physicians plus conventional resources** | **Difference (95% CI), percentage points** ^a^ | ***P* value** |
| **All participants** | 79.5 (39.1) | 69.1 (45.1) | 10.3 (1.4 to 19.3) | 0.0236 |
| **Years of Practice** |  | | | |
| Below median | 81.6 (37.7) | 66.4 (46.1) | 15.2 (2.8 to 27.6) | 0.0164 |
| Above median | 77.1 (40.6) | 73.4 (43.2) | 3.7 (-8.8 to 16.3) | 0.5621 |
| **LLM Experience** |  | | | |
| Frequent Use (weekly or more) | 80.8 (37.4) | 66.7 (47.6) | 14.1 (-0.8 to 29.0) | 0.0640 |
| Infrequent Use (less than weekly) | 78.7 (40.1) | 69.8 (44.4) | 8.9 (-2.3 to 20.1) | 0.1184 |
| **Gender** |  | | |  |
| Male | 76.3 (40.4) | 74.2 (42.8) | 2.1 (-13.9 to 18.0) | 0.8002 |
| Female | 80.9 (38.5) | 65.9 (46.3) | 15.0 (4.4 to 25.6) | 0.0057 |
| Notes:   - Abbreviation: LLM, large language model - ^a^ Estimated differences in group means were derived from a linear mixed effects model. Random effects were included for participants to account for within-participant correlations and for cases to control for case difficulty variability. Reported *P* values are two-sided. No adjustments were made for multiple comparisons. | | | | |

### Supplementary Figure 1. Study Flow Diagram

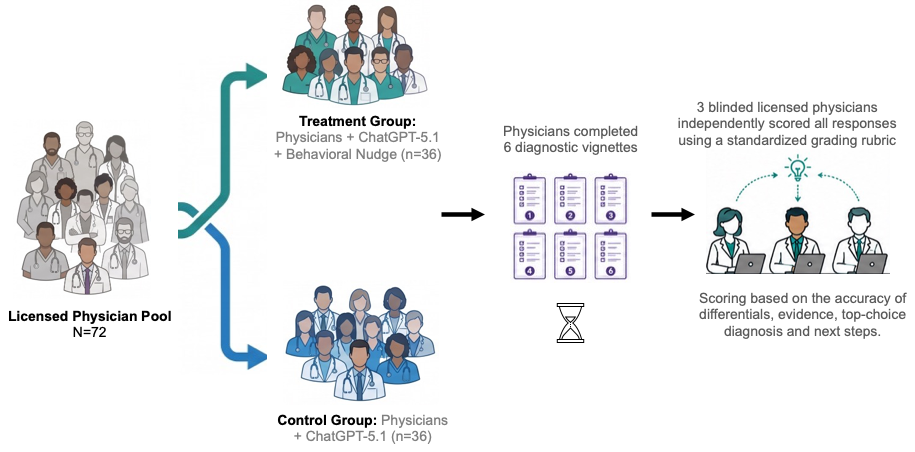

*Caption: Seventy-two licensed physicians were randomized 1:1 to one of two arms: an intervention arm receiving ChatGPT-5.1 recommendations paired with a behavioral nudge (Physicians + ChatGPT-5.1 + Behavioral Nudge; n=36) or a control arm receiving the same recommendations without the nudge (Physicians + ChatGPT-5.1; n=36). Both arms completed an identical set of six clinical vignettes, each accompanied by LLM-generated recommendations; in half of the cases, deliberate, clinically significant errors were introduced into the LLM recommendations by the physician panel. All responses were independently scored by three blinded licensed physicians using a standardized grading rubric assessing the accuracy of differential diagnoses, supporting evidence, top-choice diagnosis, and recommended next steps.*

### Supplementary Figure 2: Screenshot of the Interface Showing Clinical Vignette & Option to View ChatGPT Recommendations

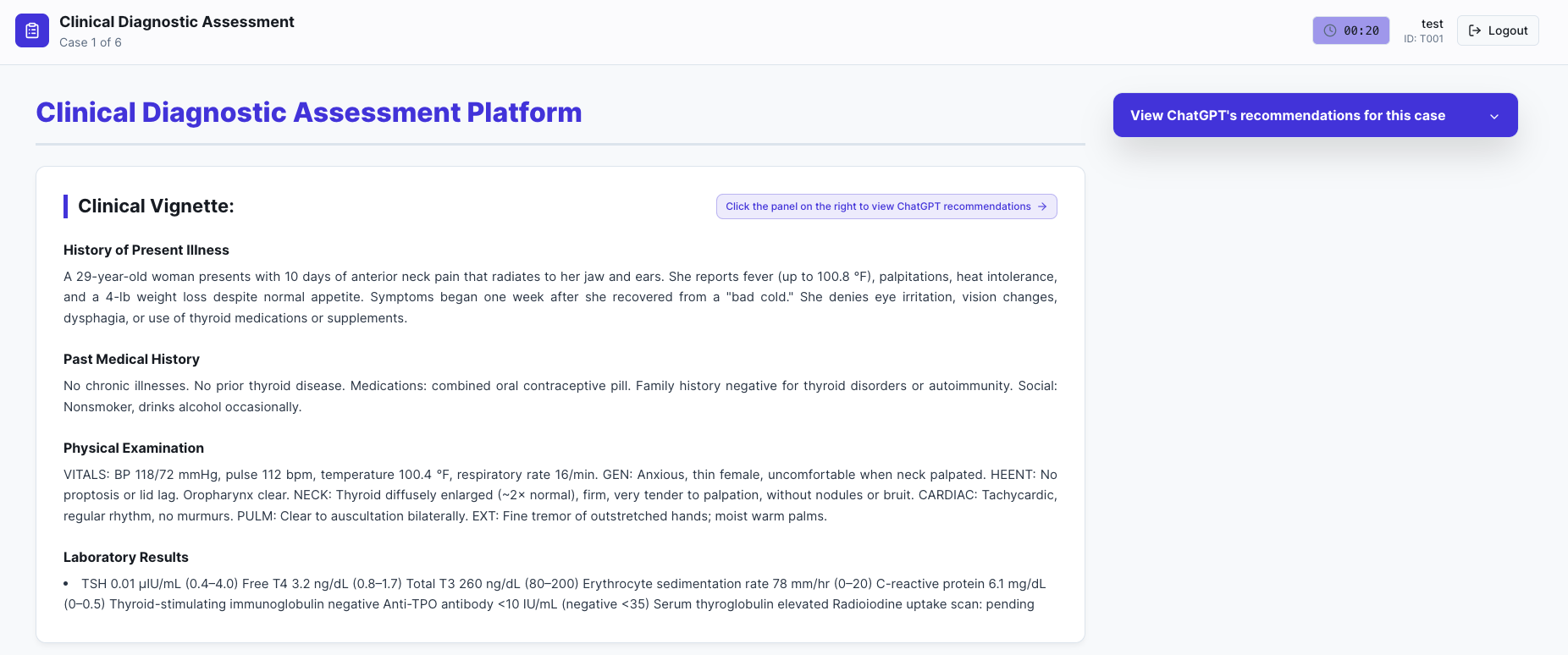

*Caption: The screenshot shows the study interface as seen by participants. ChatGPT recommendations were hidden by default and displayed only after participants clicked on the button “View ChatGPT’s recommendations for this case.”*

### Supplementary Figure 3: Screenshot of the Interface Showing Clinical Vignette & LLM Recommendations Along with a Nudge (Low Confidence Example)

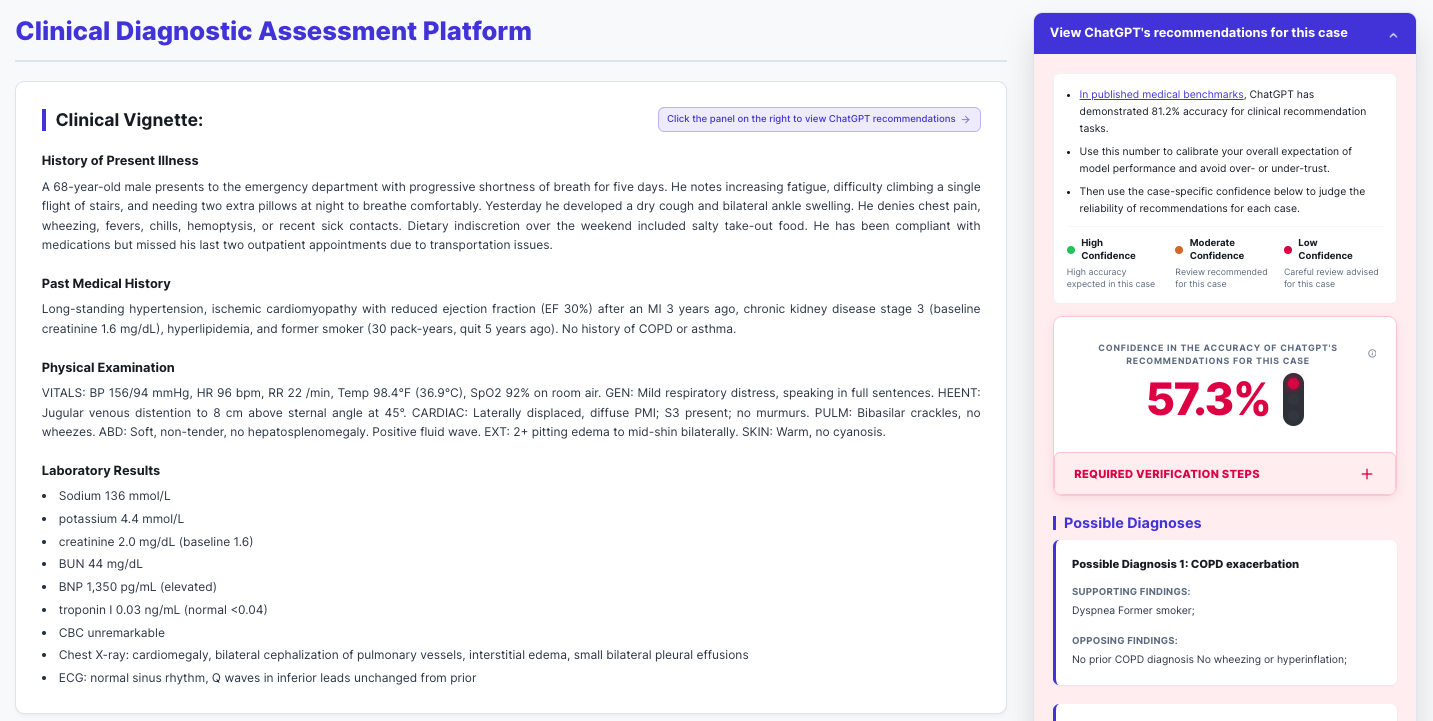

*Caption: The screenshot shows the study interface as seen by participants in the treatment group. ChatGPT recommendations are shown after the participant clicks on the “View ChatGPT’s recommendations for this case” button along with the behavioral nudge showing (i) ChatGPT’s accuracy on published medical benchamarks (i.e., 81.2% at the time, participants could click and view the link for details) and (ii) case-specific mean confidence score (i.e., 57.3%) along with a coded signal.*

### Supplementary Figure 4: Screenshot of the Nudge Interface along with the LLM Recommendations (Low Confidence Example)

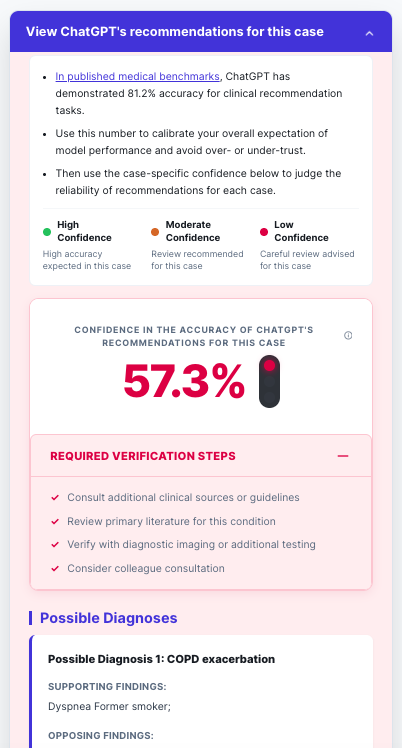

*Caption: The screenshot shows the nudge shown to participants in the treatment group. The dual-component nudge shows (i) ChatGPT’s accuracy on published medical benchamarks to calibrate overall expectations of model performance and avoid over- or under- trust and (ii) case-specific mean confidence score derived from the confidence ratings of three LLMs from distinct model families along with a coded signal and guidance on verification steps the physician should undertake (e.g., ‘consult additional clinical sources or guidelines’ and ‘review primary literacture for this condition’).*

### Supplementary Figure 5: Screenshot of the Interface Showing Clinical Vignette & LLM Recommendations Along with a Nudge (Higher Confidence Example)

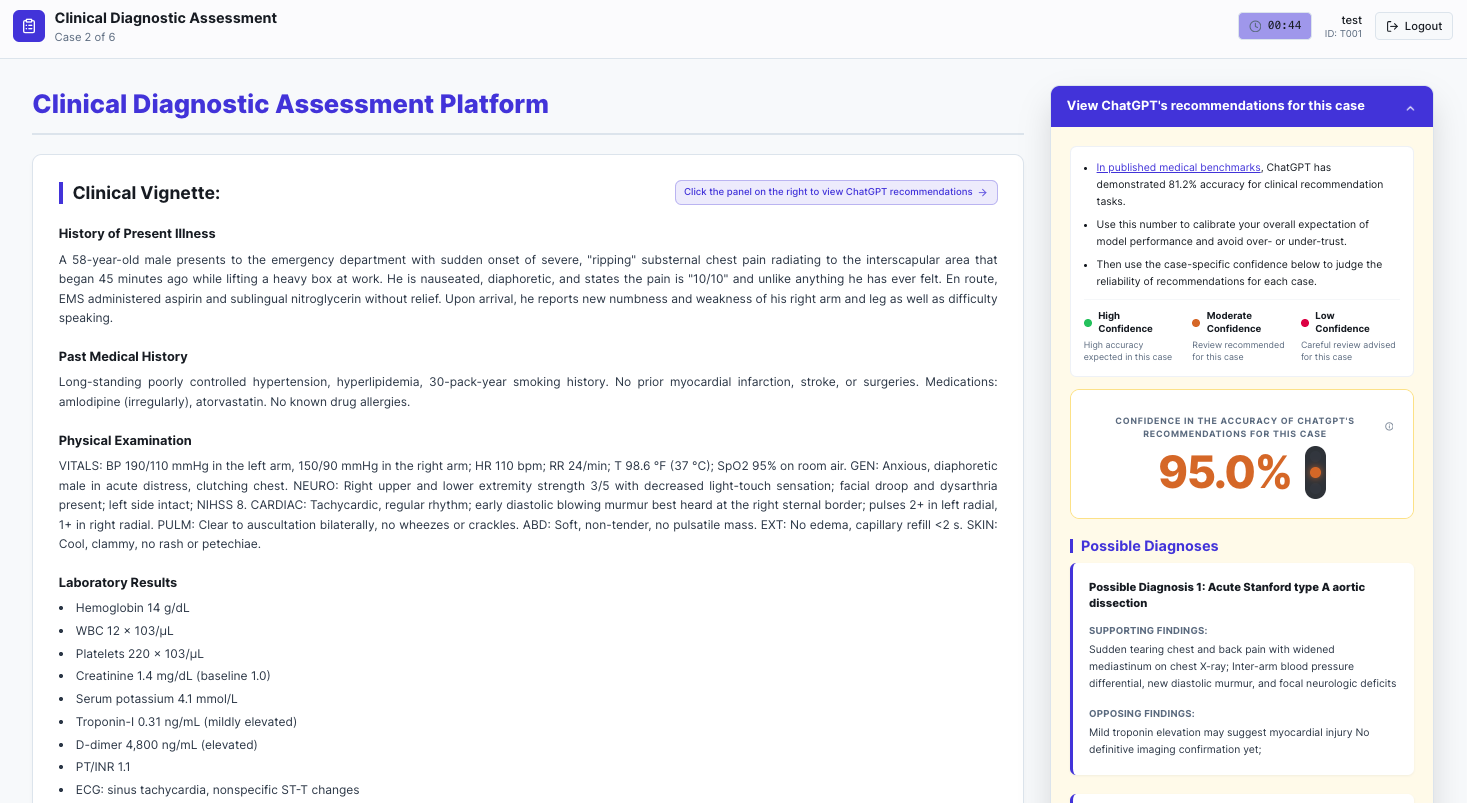

*Caption: The screenshot shows the study interface as seen by participants in the treatment group for the clinical vignette with a mean confidence rating of 95%. Standard review/oversight was recommendation.*

### Supplementary Figure 6: Screenshot of the Interface Showing Clinical Vignette & LLM Recommendations in the Control Group

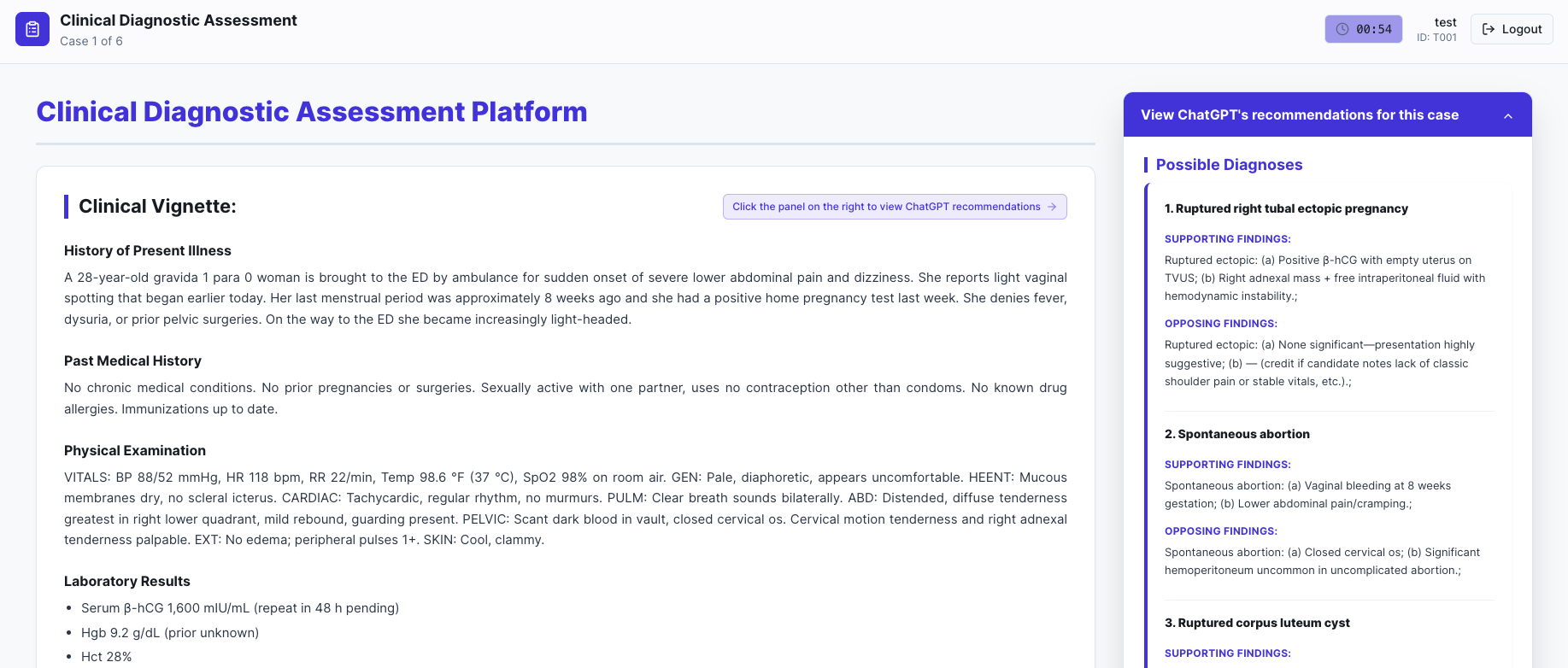

*Caption: The screenshot shows the study interface as seen by participants in the control group. No nudge was delivered to these participants.*

### Supplementary Figure 7. Statistical Validation

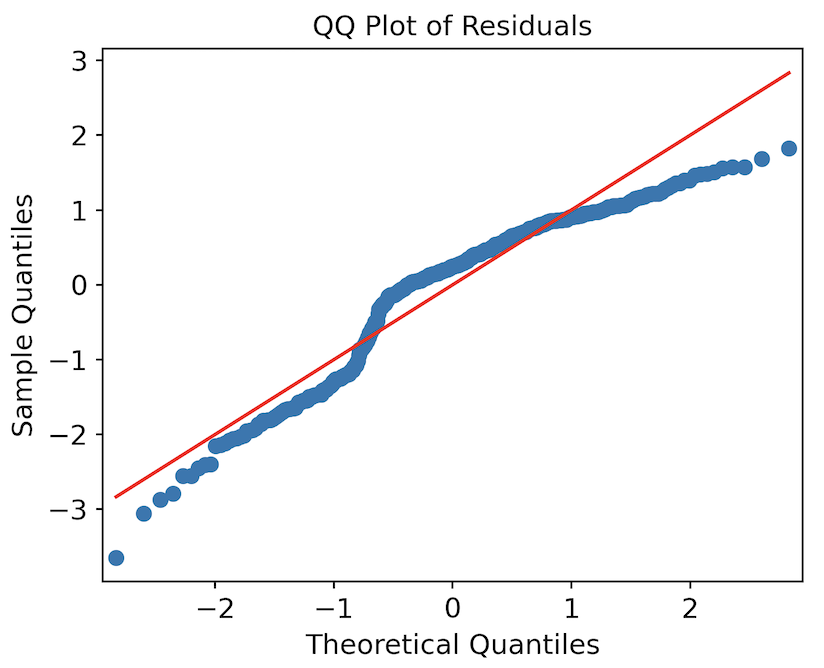

*Caption: Quantile-quantile plot of residuals from the mixed-effects model. The plot compares the distribution of model residuals (y-axis) to a theoretical normal distribution (x-axis). Points following the diagonal line indicate normality; deviations suggest departures from normal distribution. QQ plots revealed some departure from normality in the residuals, with deviations primarily in the distribution tails. However, mixed-effects models are robust to moderate violations of normality assumptions, and the observed deviations were not considered substantial enough to affect the validity of the statistical inferences.*

### Supplementary Note 1: Study Protocol & Statistical Analysis Plan

This randomized clinical trial was prospectively registered on ClinicalTrials.gov ([NCT07328815](https://clinicaltrials.gov/study/NCT07328815); registered January 9, 2026; first submitted December 26, 2025). The prespecified statistical analysis plan was uploaded to the registry on December 26, 2025.

***Study Description***

**Brief Summary**: This randomized controlled trial evaluates whether behavioral nudges can reduce automation bias, the uncritical acceptance of automated output, among AI-trained physicians using a large language model (ChatGPT-5.1) for clinical decision-making. LLM recommendations for all clinical vignettes were generated by ChatGPT-5.1, and the confidence in the accuracy of each recommendation was independently rated by three LLMs from distinct model families (Claude Sonnet 4.5, Gemini 2.5 Pro Thinking, and GPT-4o). The trial tests whether a dual-mechanism nudge, combining baseline accuracy anchoring with a case-specific signal that displays the mean of these three confidence ratings together with a color code derived from that score (red below baseline, orange above baseline but below 100%, green at 100%), reduces physicians’ uncritical acceptance of incorrect LLM recommendations.

**Condition or disease**: Diagnosis

**Intervention/treatment**: Other: Behavioral Nudge Intervention

**Phase**: Not Applicable

**Detailed Description:**

Automation bias represents a critical challenge in modern clinical practice, particularly as artificial intelligence (AI) tools become increasingly embedded in healthcare workflows. This cognitive phenomenon describes the tendency of clinicians to favor suggestions from automated decision-making systems, even when those suggestions are incorrect. As large language models (LLMs), such as ChatGPT-5.1, gain traction in medical settings, their potential to reduce errors and improve efficiency must be weighed against a significant concern: these models lack rigorous medical validation and may amplify existing cognitive biases through incorrect or misleading recommendations.

The emergence of automation bias in medical contexts reflects a complex interplay of environmental and psychological factors, including time constraints in high-volume settings, financial incentives that prioritize efficiency over thoroughness, and cognitive fatigue during extended shifts. These pressures interact with psychological mechanisms, including diffusion of responsibility, overconfidence in technological solutions, and cognitive offloading, collectively creating conditions where uncritical acceptance of AI-generated recommendations becomes more likely.

This randomized controlled trial evaluates the effectiveness of a behavioral nudge intervention designed to mitigate automation bias among physicians using LLM-generated diagnostic recommendations. The primary objective is to determine whether the intervention improves diagnostic reasoning performance scores when physicians evaluate clinical vignettes that include deliberately flawed LLM recommendations; secondary objectives assess whether physician experience level, gender, and prior LLM experience moderate the intervention’s effectiveness. The study employs a single-blind, two-arm parallel design with 1:1 randomization. To eliminate variability arising from differences in prompting skills, participants do not interact with a live LLM interface; instead, all participants use a custom-built web platform that displays clinical vignettes alongside pre-generated, fixed ChatGPT-5.1 recommendations, ensuring identical content for each vignette.

All participants will evaluate six clinical vignettes during a single, proctored session lasting approximately 75 minutes. Three vignettes will contain deliberately introduced clinical reasoning flaws in the LLM recommendations, while three will contain correct recommendations. Vignettes will be presented in randomized order to prevent pattern detection.

Control arm participants will evaluate clinical vignettes with LLM diagnostic recommendations generated by ChatGPT presented in standard, neutral text format without additional contextual information. Intervention arm participants will evaluate the same vignettes alongside a behavioral nudge. This intervention consists of two synchronized cognitive cues: (1) an anchoring cue displaying ChatGPT's baseline diagnostic accuracy on standard medical datasets at the top of the interface panel, explicitly anchoring expectations to the model's fallibility, and (2) a selective attention cue displaying the LLM recommendation alongside a color-coded confidence signal generated through an ensemble assessment: three independent state-of-the-art LLMs (Claude Sonnet 4.5, Gemini 2.5 Pro Thinking, and GPT-4o) each provide confidence ratings for the recommendation, and the mean confidence determines the signal color to mitigate single-model miscalibration.

The color-coded confidence signals are categorized into three distinct levels based on the ensemble's mean confidence relative to baseline diagnostic accuracy. Red signals are triggered when the mean confidence falls below ChatGPT's established baseline accuracy, explicitly flagging high-uncertainty cases that demand heightened critical scrutiny. Orange signals indicate that while the mean confidence exceeds the baseline average, it remains below 100%, signaling the need for continued clinical vigilance and the avoidance of complacency. Finally, green signals are reserved for instances of 100% ensemble consensus; however, even at this level of confidence, standard AI safety warnings remain present to guard against over-reliance on the system's output.

Participants will be presented with six clinical vignettes specifically designed to measure automation bias, sourced and modified from real cases representing a range of diagnostic difficulty and common medical specialties. Each vignette follows a standardized format including chief complaint, history of present illness, relevant past medical/social/family history, physical examination findings, and initial laboratory results.

The primary outcome is the Diagnostic Reasoning Accuracy Score, a composite percentage score based on a structured rubric evaluating: quality of differential diagnoses, supporting findings, opposing findings, final diagnosis accuracy, and appropriateness of next steps. The secondary outcome is the Top Choice Diagnosis Accuracy Score. All responses will be evaluated by blinded reviewers using the assessment rubric.

***Study Design***

Study Type: Interventional (Clinical Trial)

Estimated Enrollment: 50 participants

Allocation: Randomized. Participants will be assigned in a 1:1 ratio using a computer-generated random number sequence.

Intervention Model: Parallel Assignment

Intervention Model Description: The trial will be designed as a randomized, two-arm, single-blind parallel group study.

Masking: Single (Outcomes Assessor)

Masking Description: The grading of responses will be performed by assessors blinded to participant identity and treatment assignment.

Primary Purpose: Diagnostic

Official Title: Mitigating Automation Bias in Physician-LLM Diagnostic Reasoning Using Behavioral Nudges

Actual Study Start Date: January 17, 2026

Actual Primary Completion Date: May 25, 2026

Actual Study Completion Date: May 26, 2026

***Arms and Interventions***

| **Arm** | **Intervention/Treatment** |
| --- | --- |
| **Active Comparator: ChatGPT Recommendations alongside a Behavioral Nudge**  Participants will evaluate six clinical vignettes. During the trial, they will have access to clinical recommendations from a specific, commercially available LLM (ChatGPT) in addition to conventional diagnostic resources. LLM recommendations for three vignettes will contain deliberately flawed diagnostic information, and for three vignettes they will contain accurate recommendations. The cases will be presented in random order. Participants in this arm will receive a behavioral nudge embedded in the LLM recommendations interface that presents two synchronized cognitive cues when the LLM panel is expanded: (1) an anchoring cue displaying ChatGPT’s baseline diagnostic accuracy on standard medical datasets at the top of the panel to set realistic expectations, and (2) a selective attention cue located immediately below, which shows the LLM recommendation alongside a case-specific, color-coded confidence signal. | **Other: Behavioral Nudge Intervention**  Participants in the treatment group will receive a behavioral nudge intervention embedded in the LLM recommendations interface that presents two synchronized cognitive cues when the LLM panel is expanded: (1) an anchoring cue displaying ChatGPT’s baseline diagnostic accuracy on standard medical datasets at the top of the panel to set realistic expectations before viewing the specific recommendation, and (2) a selective attention cue located immediately below, which shows the LLM recommendation alongside a case-specific, color-coded confidence signal. This signal is categorized as red when the mean ensemble confidence falls below the established baseline accuracy, flagging high-uncertainty cases that demand critical evaluation; orange when confidence meets or exceeds the baseline but remains below 100%, intended to prevent complacency and maintain active clinical scrutiny; and green for a 100% ensemble consensus, though standard cautionary warnings still apply to guard against over-reliance. |
| **No Intervention: ChatGPT Recommendations without a Behavioral Nudge**  Participants will evaluate six clinical vignettes. During the trial, they will have access to clinical recommendations from a specific, commercially available LLM (ChatGPT) in addition to conventional diagnostic resources. LLM recommendations for three vignettes will contain deliberately flawed diagnostic information. The cases will be presented in random order. Participants in this arm will not receive any behavioral nudge. |  |

***Outcome Measures***

**Primary Outcome Measures:**

Diagnostic reasoning accuracy score [Time Frame: Assessed at a single time point for each case, during the scheduled diagnostic reasoning evaluation session, which takes place between 0-5 days after participant enrollment.]

The primary outcome will be the percent correct for each case, ranging from 0 to 100%, where higher scores indicate better diagnostic performance. For each case, participants will be asked for their three leading diagnoses, findings that support each diagnosis, and findings that oppose each diagnosis. For each plausible diagnosis, participants will receive 1 point. Findings supporting the diagnosis and findings opposing the diagnosis will also be graded based on correctness, with 1 point for each correct response. Participants will then be asked to name their top diagnosis they believe is most likely, earning 9 points for a reasonable response and 18 points for the most accurate response. Finally, participants will be asked to name up to 3 next steps to further evaluate the patient with 0.5 points awarded for a partially correct response and 1 point for a completely correct response. The primary outcome will be compared at the case-level between the randomized groups.

**Secondary Outcome Measures:**

Top choice diagnosis accuracy score [Time Frame: Assessed at a single time point for each case, during the scheduled diagnostic reasoning evaluation session, which takes place between 0-5 days after participant enrollment.]

The secondary outcome will measure participants’ performance in identifying the most likely diagnosis for each clinical vignette. After evaluating each case, participants will select their single most likely diagnosis, which will be scored on a pre-specified Three-Tier Diagnostic Accuracy Scale: 18 points for the most accurate diagnosis, 9 points for a clinically reasonable alternative, and 0 points for an incorrect diagnosis. For each participant, a Top Choice Diagnosis Accuracy Score is calculated as (total points earned ÷ maximum possible points) × 100, yielding a 0-100 % range in which higher scores indicate greater diagnostic accuracy. This percentage score will be compared at the case-level between randomized groups to quantify the impact of automation bias on diagnostic decision-making.

***Eligibility Criteria***

Ages Eligible for Study: All

Sexes Eligible for Study: All

Accepts Healthy Volunteers: Yes

**Criteria**

Inclusion Criteria:

- Completed Bachelor of Medicine, Bachelor of Surgery (MBBS) Exam. The equivalent degree of MBBS in US and Canada is called Doctor of Medicine (MD).
- Full or Provisionally Registered Medical Practitioners with the Pakistan Medical and Dental Council (PMDC).
- Participants must have completed a structured training program on the use of ChatGPT (or a comparable large language model), totaling at least 10 hours of instruction. The program must include hands-on practice related to the LLM’s key aspects, specifically prompt engineering and content evaluation.

Exclusion Criteria:

- Any other Registered Medical Practitioners (Full or Provisional) with PMDC (e.g., Professionals with Bachelor of Dental Surgery or BDS).

**Potential Risks and Harms**

This study poses minimal risk to participants. Some participants may experience mild discomfort or frustration, particularly those in the treatment group who may find clinical vignettes challenging when the accompanying LLM recommendations contain deliberate errors.

**Safeguards for Addressing Risks or Harms to Subjects**

To minimize potential discomfort, participants will receive comprehensive instructions and ongoing support throughout the study. Participants will be explicitly informed that individual performance is not being evaluated, and that data will be analyzed for aggregate trends only. All participants will be advised of their right to withdraw from the study at any time without penalty or explanation. Research staff will monitor for signs of distress and provide appropriate support as needed.

**Power Analysis:**

The minimum target sample size of 50 participants (25 participants per arm) was predetermined based on a prior study.^1^ The power analysis, conducted using Python version 3.11.9 (Python Software Foundation), employed the statsmodels.stats.power module from statsmodels version 0.14.4 (Statsmodels Developers) and indicated that a total sample of 200 to 250 completed cases (approximately 4 to 5 cases per participant) would provide at least 80% power to detect an 8-percentage-point mean difference in diagnostic reasoning scores, assuming a two-sided α of .05. The analysis employed mixed-effects models suitable for cluster-randomized designs, considering an intraclass correlation coefficient (ICC) ranging from 0.05 to 0.15 and a standard deviation of 16.2%.

**Statistical Analysis:**

Descriptive analysis will be performed by comparing participant characteristics between the treatment and control groups. Categorical variables will be reported using counts and proportions; continuous variables as means and standard deviations, or medians with interquartile ranges if non-normally distributed.

The primary outcome analysis will be conducted at the case level, with clustering by participant under an intention-to-treat framework. The primary analysis will include cases with completed responses only. We will first summarize the mean and standard deviation of scores (standardized on a 0-100 scale) for both groups. To evaluate the impact of the behavioral nudge on automation bias, we will apply linear mixed-effects models, including a random effect for the participant to account for potential within-participant correlation across cases, and a random effect for cases to account for varying case difficulty. The primary analysis will include the following as covariates for adjustment: experience in LLM use, gender, and years of practice post MBBS. Subgroup analyses will be performed based on experience with LLMs, gender, and years of experience post-MBBS.

While the study design minimizes the potential for missing data, if missing data does occur, multiple imputation will be considered under the intention-to-treat framework for the primary analysis. No interim analyses are planned given the small sample size.

All statistical analyses will be performed using Python software, version 3.11.12 (Python Software Foundation) with pandas for data manipulation and statsmodels version 0.14.4 for mixed-effects modeling. Statistical significance will be based on a *p* value.

***Contacts and Locations***

Locations

Pakistan, Lahore

Lahore University of Management Sciences

Lahore, Punjab 54792

Sponsors and Collaborators

Lahore University of Management Sciences

Investigators

Principal Investigator: Ihsan Ayyub Qazi, PhD, Lahore University of Management Sciences

Principal Investigator: Muhammad Hamad Alizai, PhD, Lahore University of Management Sciences

Principal Investigator: Muhammad Asad Ullah Khawaja, MBBS, King Edward Medical University

Principal Investigator: Ali Zafar Sheikh, MBBS, Lahore General Hospital

Principal Investigator: Muhammad Junaid Akhtar, MBBS, Children's Hospital, Lahore

***References***

1. Goh E, Gallo R, Hom J, et al. Large Language Model Influence on Diagnostic Reasoning: A Randomized Clinical Trial. *JAMA Netw Open.* 2024;7(10):e2440969. doi:10.1001/jamanetworkopen.2024.40969

### Supplementary Note 2: Model Specification

Our study employed a linear mixed effects model with 72 participants, each of whom completed 6 identical clinical cases. This design requires modeling three variance components: between-participant ($\sigma_{a}^{2}$), between-case ($\sigma_{c}^{2}$), and residual ($\sigma_{e}^{2}$).

The model specification is:

*y*ij = β0 + β1*X*1i + β2*X*2i + β3*X*3i + β4*X*4i + *a*i + *c*j + *e*ij

where:

*y*ij is the average score (percentage) for participant *i* on case *j*

*X*1i is the treatment indicator (1 if treatment, 0 if control)

*X*2i is gender (1 if male, 0 otherwise)

*X*3i is LLM usage (1 if frequent , 0 otherwise)

*X*4i is experience (in years)

*a*i ~ N(0, $\sigma_{a}^{2}$),) is the participant random intercept

*c*j ~ N(0, $\sigma_{c}^{2}$) is the case random effect

*e*ij ~ N(0, $\sigma_{e}^{2}$) is the residual error

The estimated variance components were:

Between-participant: $\sigma_{a}^{2}$ = 67.56

Between-case: $\sigma_{c}^{2}$ = 426.87

Residual: $\sigma_{e}^{2}$ = 218.04

The Intraclass Correlation Coefficient (ICC) for participant clustering is:

ICC = $\sigma_{a}^{2}$ / ($\sigma_{a}^{2}$ + $\sigma_{c}^{2}$ + $\sigma_{e}^{2}$) = 67.557 / (67.56 + 426.87 + 218.04) = 67.56 / 712.47 = 0.095

The ICC of 0.095 falls within our a priori power calculation assumptions (ICC: 0.05-0.15), confirming adequate statistical power.

**Table 1. Mixed Linear Model Regression Results**

| **Parameter** | **Coefficient** | **SE** | **z** | ***P*-value** | **95% CI** |
| --- | --- | --- | --- | --- | --- |
| Intercept | 76.85 | 3.12 | 24.64 | <0.001 | 70.74 to 82.96 |
| Treatment | 7.64 | 3.19 | 2.40 | 0.016 | 1.40 to 13.89 |
| Male (vs Female, reference) | -0.09 | 3.33 | -0.03 | 0.979 | -6.61 to 6.43 |
| Infrequent LLM use (vs Frequent, reference) | -1.89 | 3.49 | -0.54 | 0.588 | -8.73 to 4.95 |
| Years in Practice | -0.10 | 0.20 | -0.50 | 0.621 | -0.50 to 0.30 |

*Caption: Table 1 presents the complete model results, including coefficient estimates, standard errors, z-statistics, p-values and 95% confidence intervals.*

*Note.* SE = standard error. Model estimated with 72 participants (groups) and 432 observations.
